# COVID Seroprevalence, Symptoms and Mortality During the First Wave of SARS-CoV-2 in Canada

**DOI:** 10.1101/2021.03.04.21252540

**Authors:** Action to beat coronavirus/Action pour battre le coronavirus (Ab-C) Study Investigators, Prabhat Jha

## Abstract

**Background:** Efforts to stem Canada’s SARS-CoV-2 pandemic can benefit from direct understanding of the prevalence, infection fatality rates (IFRs), and information on asymptomatic infection.

**Methods:** We surveyed a representative sample of 19,994 adult Canadians about COVID symptoms and analyzed IgG antibodies against SARS-CoV-2 from self-collected dried blood spots (DBS) in 8,967 adults. A sensitive and specific chemiluminescence ELISA detected IgG to the spike trimer. We compared seroprevalence to deaths to establish IFRs and used mortality data to estimate infection levels in nursing home residents.

**Results:** The best estimate (high specificity) of adult seroprevalence nationally is 1.7%, but as high as 3.5% (high sensitivity) depending on assay cut-offs. The highest prevalence was in Ontario (2.4-3.9%) and in younger adults aged 18-39 years (2.5-4.4%). Based on mortality, we estimated 13-17% of nursing home residents became infected. The first viral wave infected 0.54-1.08 million adult Canadians, half of whom were <40 years old. The IFR outside nursing homes was 0.20-0.40%, but the COVID mortality rate in nursing home residents was >70 times higher than that in comparably-aged adults living in the community. Seropositivity correlated with COVID symptoms, particularly during March. Asymptomatic adults constituted about a quarter of definite seropositives, with a greater proportion in the elderly.

**Interpretation:** Canada had relatively low infection prevalence and low IFRs in the community, but not in nursing homes, during the first viral wave. Self-collected DBS for antibody testing is a practicable strategy to monitor the ongoing second viral wave and, eventually, vaccine-induced immunity among Canadian adults.

## Main Text

About 9,000 Canadians, most of them nursing home residents, died from COVID-19 (COVID) during the first viral wave from March to July 2020.^1^ The mortality rate is low compared with rates in the United States and most of Europe,^2^ but like other countries, Canada is experiencing a second wave in late 2020/early 2021, with cumulative confirmed cases exceeding 800,000 and rising mortality.

Here, we report results from the Action to Beat Coronavirus (Ab-C) seroprevalence study in a reasonably representative sample of Canadians, with self-reported COVID symptoms from over 19,000 adults,^3^ IgG antibody testing for SARS-CoV-2 from about 9,000, and national and provincial mortality data. This report is the first of a planned series of periodic national assessments to establish cumulative prevalence at each point, incidence, the relationship of age-specific mortality to SARS-CoV-2 seroprevalence and information on asymptomatic infections. Collectively, this information^4^ will help track the effectiveness of vaccination and other measures in key populations and geographies of Canada.

## Methods

### Participant Recruitment

We invited 44,270 members of the Angus Reid Forum (ARF),^5^ an established nationwide panel of Canadian adults (used for opinion polling), to take an online survey on symptoms associated with SARS-CoV-2 and testing histories. The first phase from May to June selected adults stratified by age, sex, education, and region, by census metropolitan area to match the national demographic distribution, with oversampling of adults age 60 years or older. Given concern about some geographic areas that reported increases in COVID cases during August-September (early in the second wave), we later recruited Forum panel members who lived in 17 pre-defined public health regions (out of 93 in Canada) who had not already enrolled in the study. The 17 high-burden areas had have higher infection levels, based on a regression analysis of SARS-CoV-2 case counts (Appendix Figure 2). At the end of the online poll, respondents indicated their willingness to self-collect a dried blood spot (DBS) sample from a finger prick, and consenters were sent a DBS collection kit. Participants who responded to the online poll in May and June provided DBS samples between June and August, and those who completed the poll in August and September provided DBS samples in September. Participants were not compensated financially by the study for completing the poll, but earn modest redeemable points for participation in the ARF. Appendix section 1 illustrates the study recruitment and flow, additional details of the ARF, and the regression analyses used to select high-burden areas.

### Symptoms and SARS-CoV-2 Testing

The online polls solicited a brief medical history, self-reported COVID symptoms by the month when they first occurred, and experience with COVID testing.^3^ Based on the published literature, we defined “COVID symptom-positive” as a combination of fever plus any of difficulty breathing, dry cough, loss of smell, or “COVID toe.” The same questions were asked about other household members. We asked participants if they had had a PCR test for SARS-CoV-2 or were awaiting one. Additional questions were asked about hypertension, diabetes, self-reported height and weight (to calculate body mass index [BMI]), current or past smoking, and other exposures (Appendix Table 1).^6^

### IgG Serology

We asked DBS participants to collect five small circles of blood on a special bar-coded filter paper, dry the sample for at least two hours, place it in a Mylar pouch with a desiccant (inside a second protective envelope), and return it to us at St. Michael’s Hospital in Toronto, postage prepaid. Mailing time across Canada ranged from 3 to 6 days. Upon arrival, samples were scanned, catalogued, and stored at 4°C in larger boxes with additional desiccant, and monitored for humidity levels (kept <20%). We conducted quality control by examining completeness of all five circles, with only 34 having inadequate dried blood for analyses. The Network Biology Collaborative Centre at Sinai Health, Toronto, conducted high-throughput analyses using a highly sensitive chemiluminescence-based ELISA targeting the spike protein (as a trimer). The sensitivity of the IgG spike assay on serum has been demonstrated to be ≥94%, with specificity of >99% on serum or plasma samples.^7^ Using this assay, we have shown that IgGs to the spike trimer can persist for at least 115 days in symptomatic individuals.^7^ Pilot studies with samples acquired locally or through the National Microbiology Laboratory of Canada established that eluents from contrived blood spots yielded results highly correlated to those of liquid plasma (data not shown). We used a pool of SARS-CoV-2 negative serum, human IgG serum and blanks as controls, and negative DBS controls in the batches. Appendix Figures 3-7 provides details of lab protocol, normalization, and reproducibility procedures.

### COVID Mortality

We collected age, sex, and location (nursing home or not) for COVID deaths (defined by the World Health Organization [WHO] as death codes U07.1 and U07.2) from Statistics Canada and from provincial data sources. In Canada, excess mortality recorded in 2020 was nearly all due to COVID.^1,8^ We modified the Statistics Canada definitions of long-term care residents to include residents living in mixed care and retirement homes.^9^ We conducted Bayesian analyses of the distribution of these deaths by age, sex, and nursing home or other setting assuming prior distributions for all parameters in the model and Poisson distribution for the number of deaths in nursing homes and otherwise (Appendix). From these and the national population and death totals for each age group (20-39, 40-59, 60-69, and 70+), we estimated infection fatality rates (IFRs) as deaths/prevalence of SARS-CoV-2 from the serology per 100,000 infections. We treated the approximate 211,000 nursing home population in 2019 as its own age/risk group,^10^ noting that the vast majority of residents were aged 80 or older.

### Data Analysis

The main calculation is of IgG (spike trimer) antibody prevalence, categorizing results as definite (higher specificity: ≥3 standard deviations from mean of samples previously shown to be negative as both plasma and as contrived dried blood spots), possible (higher sensitivity: same procedure but using presumed negatives within the study as the comparator), or negative. We standardized prevalences for age and education profiles of the 2016 census. Documenting nursing home COVID deaths is central to understanding IFRs in the whole population^11^ (Appendix Table 4). Given the lack of any reliable PCR or antibody testing in nursing homes, we used a mortality-based estimate of their prevalence of infection.^12^ We drew on estimated IFRs from a demographic analysis of deaths in nursing homes in France,^11^ which are similar demographically to nursing homes in Canada.^13^ We used logistic regression to examine the individual predictors of IgG antibody status, symptoms, or asymptomatic infections, using Stata 16.^14^ The Ab-C study is approved by the Unity Health Toronto Ethics Review Board.

## Results

Of the 44,270 invited Forum panel members, 19,994 completed the online survey (14,641 in Phase 1 during May-June, and 5,353 in Phase 2 during August-September). Table 1 provides the demographics and health characteristics of those completing the poll, alongside a comparison of the distribution of these characteristics in the Canadian population. The older age distribution of the respondents in Phase 1 versus the Census is a result of intentionally oversampling individuals age 60 and older. Other demographic characteristics of the respondents in the Ab-C study were similar to those from the 2016 Census,^3,15-16^ with the exception of an under-representation of adults without a completed college education (24% in our sample versus 46% in Canada). Hence, all subsequent prevalence estimates adjusted for education. Despite this, participants providing either polling results or DBS samples in both phases were similar to the national prevalences of obesity, current smoking, diabetes, and hypertension (Tables 1 and 3).^17-20^

**Table 1.** Representativeness of online poll, dried blood spot sample, and SARS CoV-2 seroprevalence in Canada, overall 2020

|  |  |  |  |  |  |  |
| --- | --- | --- | --- | --- | --- | --- |
| Comparison group | Census 2016 distribution | Survey sample | Dried blood spot sample | No of definite/<br>possible | Seroprevalence |  |
|  |  |  |  |  | Definite | Definite or possible |
| **Both Phases** |  | 19,994 (100%) | 8,967 (100%) | 168 / 180 | 1.87% | 3.91% |
| Phase 1 (May-July) |  | 14,641 (73%) | 7,068 (79%) | 120 / 128 | 1.70% | 3.53% |
| Phase 2 (Aug-Sept) |  | 5,353 (27%) | 1,899 (21%) | 48 / 52 | 2.54% | 5.44% |
| Higher risk regions in Phase 1 (May-July) |  | 5,060 (25%) | 2,458 (27%) | 56 / 52 | 1.77% | 3.53% |
| Higher risk regions in Phase 2 (Aug-Sept) |  | 2,481 (12%) | 891 (10%) | 26 / 24 | 2.38% | 3.69% |
| **Results by province, age and sex for Phase 1** |  |  |  |  |  |  |
| **Province** |  |  |  |  |  |  |
| Ontario | 38% | 5,680 (39%) | 3,007 (43%) | 71 / 47 | 2.35% | 3.91% |
| British Columbia & Yukon | 14% | 2,112 (14%) | 1,049 (15%) | 10 / 22 | 0.90% | 2.95% |
| Quebec | 23% | 2,900 (20%) | 1,262 (18%) | 18 / 26 | 1.56% | 3.57% |
| Prairies & Northwest Territories | 19% | 2,756 (19%) | 1,214 (17%) | 14 / 26 | 1.14% | 3.41% |
| Atlantic provinces | 7% | 1,193 (8%) | 536 (8%) | 7 / 7 | 1.32% | 2.78% |
| **Age groups** |  |  |  |  |  |  |
| 18 to 39 years | 49% | 4,143 (28%) | 1,838 (26%) | 46 / 34 | 2.48% | 4.36% |
| 40 to 59 years | 28% | 4,309 (29%) | 2,125 (30%) | 26 / 33 | 1.35% | 2.88% |
| 60 to 69 years | 12% | 4,088 (28%) | 2,095 (30%) | 41 / 40 | 1.92% | 3.90% |
| 70 years or older | 11% | 2,101 (14%) | 1,010 (14%) | 7 / 7 | 0.69% | 2.77% |
| **Sex** |  |  |  |  |  |  |
| Male | 49% | 7,001 (48%) | 2,995 (42%) | 49 / 58 | 1.68% | 3.60% |
| Female | 51% | 7,565 (52%) | 4,053 (57%) | 71 / 70 | 1.73% | 3.51% |

Notes: 165 survey and 47 DBS participants preferred to self-describe their sex and in these 2 definite positives are not shown

Overall, there were 168 definite and 180 possible seropositives in the entire study; 120 and 128, respectively, were in Phase 1 (Table 1). The national and regional prevalence estimates are based on Phase 1 results alone, as Phase 2 participants were selected from areas suspected to have higher virus activity. The overall seroprevalence for definite seropositives, standardized for age and education levels was 1.7% (95% CI: 1.4-2.0%); adding possible seropositives, it was 3.5% (95% CI: 3.1-4.0%). Diagnostic accuracy (the two-fold difference between definite and possible results) rather than random variation is the main source of uncertainty. Hence, our main results present the standardized prevalence as ranges of definite (high specificity) or definite plus possible seropositivity (high sensitivity). Seroprevalence peaked at ages 18-39, fell at older ages, and was similar in men and women. Ontario had the highest overall adult seroprevalence (2.4-3.9%), followed by Quebec (1.6-3.6%), and British Columbia and Yukon had the lowest (0.9-3.0%), although the low absolute number of seropositives resulted in overlapping confidence intervals.

Because the Ab-C Phase 1 sampling was broadly representative of the Canadian adult population, we were able to make plausible estimates of the total number of Canadians seropositive to SARS-CoV-2, and compare seroprevalence to deaths to derive the IFR (Table 2). The overall education-adjusted seroprevalence was 2-4% at ages 20-69 years, and lower at ages 70 or older. We estimate that 0.54-1.08 million adult Canadians had SARS-CoV-2 antibodies through September 1, 2020, nearly half of them young adults aged 20-39. The COVID death rates per population were low at ages 20-69 years, but rose sharply with age, peaking at ages 80 or older. In Canada, fully 7,009 of the 9,045 (77%) recorded COVID deaths as of September 1 occurred in nursing homes.^1,10^ The death rate in nursing homes was more than 70 times greater than that among adults age 80 or older living outside of nursing homes. The IFRs were low for people <70 years old, rising to 1.1-4.4% at ages 70 or older. The mortality-based prevalence of SARS-CoV-2 infection suggested that 13-17% of Canadian nursing home residents were infected—4-8 times higher than the rate in the general population—translating to an estimate of 27,000-37,000 seropositive residents. Sensitivity analyses using lower and higher IFRs suggested prevalences ranging from 9-32% among nursing home residents (Appendix).

**Table 2.** Age-specific distribution of COVID deaths, SARS-CoV-2 infection and infection fatality rates in Canada, 2020

|  |  |  |  |  |  |  |  |  |  |
| --- | --- | --- | --- | --- | --- | --- | --- | --- | --- |
| Age group (years) | No deaths COVID deaths by Aug 31 | Population (millions) | COVID mortality rate per 100,000 | Seroprevalence |  | Nos of infected in thousands |  | Infection fatality rate * |  |
|  |  |  |  | Definite | Definite or possible | Lower | Upper | Lower | Upper |
| 20-39 | 24 | 10.4 | 0.23 | 2.5% | 4.4% | 258 | 454 | 0.005% | 0.009% |
| 40-59 | 223 | 9.9 | 2.25 | 1.4% | 2.9% | 134 | 286 | 0.08% | 0.17% |
| 60-69 | 490 | 4.7 | 10.36 | 1.9% | 3.9% | 91 | 184 | 0.27% | 0.54% |
| 70 or older† | 1,299 | 4.3 | 30.59 | 0.7% | 2.8% | 29 | 118 | 1.10% | 4.43% |
| Nursing home populations† | 7,009 | 0.2 | 3318.39 | (12.9%-17.4%) |  | 27 | 37 | 19.06% | 25.75% |
| Total adult population‡ | 9,045 | 29.5 | 30.63 | 1.7% | 3.5% | 539 | 1,079 | 0.20%§ | 0.40%§ |

\* By comparison, a global review found average infect fatality rates of 0.04%, 0.36%, 0.77%, and 10.73% at ages 20-39, 40-59, 60-69 and 70+, respectively<sup>11</sup>
† For 70-79 years and for 80+ years, the COVID deaths, population (millions), mortality rate per 100,000 were: 679, 3.0, and 22.60; and 620, 1.5, and 42.69, respectively.
‡ Mortality-derived prevalence. See methods and Supplementary Appendix section 4 for details.
§ Infection fatality rate in total population, excluding nursing home populations

The analyses of determinants of seropositivity were similar in Phase 1 and 2 and are combined. 41 (24%) of the definite and 68 (38%) of the possible seropositives reported none of the study definition symptoms, meaning that 31% of those infected were asymptomatic. Asymptomatic proportions using the highly sensitive cut-offs were higher in those aged 70 or older (62%) or aged 60-69 years (41%) versus 18-49 years (20%) but lower among Indigenous populations (17%) than among English or European ethnicity (35%), with similar results in multivariate analyses (Appendix Tables 1-2). COVID symptoms were notably more prevalent among definite seropositives than among possible seropositives. Seroprevalence among those who were COVID symptom-positive was notably higher (8.7-17.8%) than in those without COVID symptoms (1.2-2.6%). The peak seroprevalence among those with symptoms was in those reporting symptom onset in March (17.7-36.4%; Table 3, Appendix Tables 2-3, Appendix Figures 8-9).

**Table 3.**
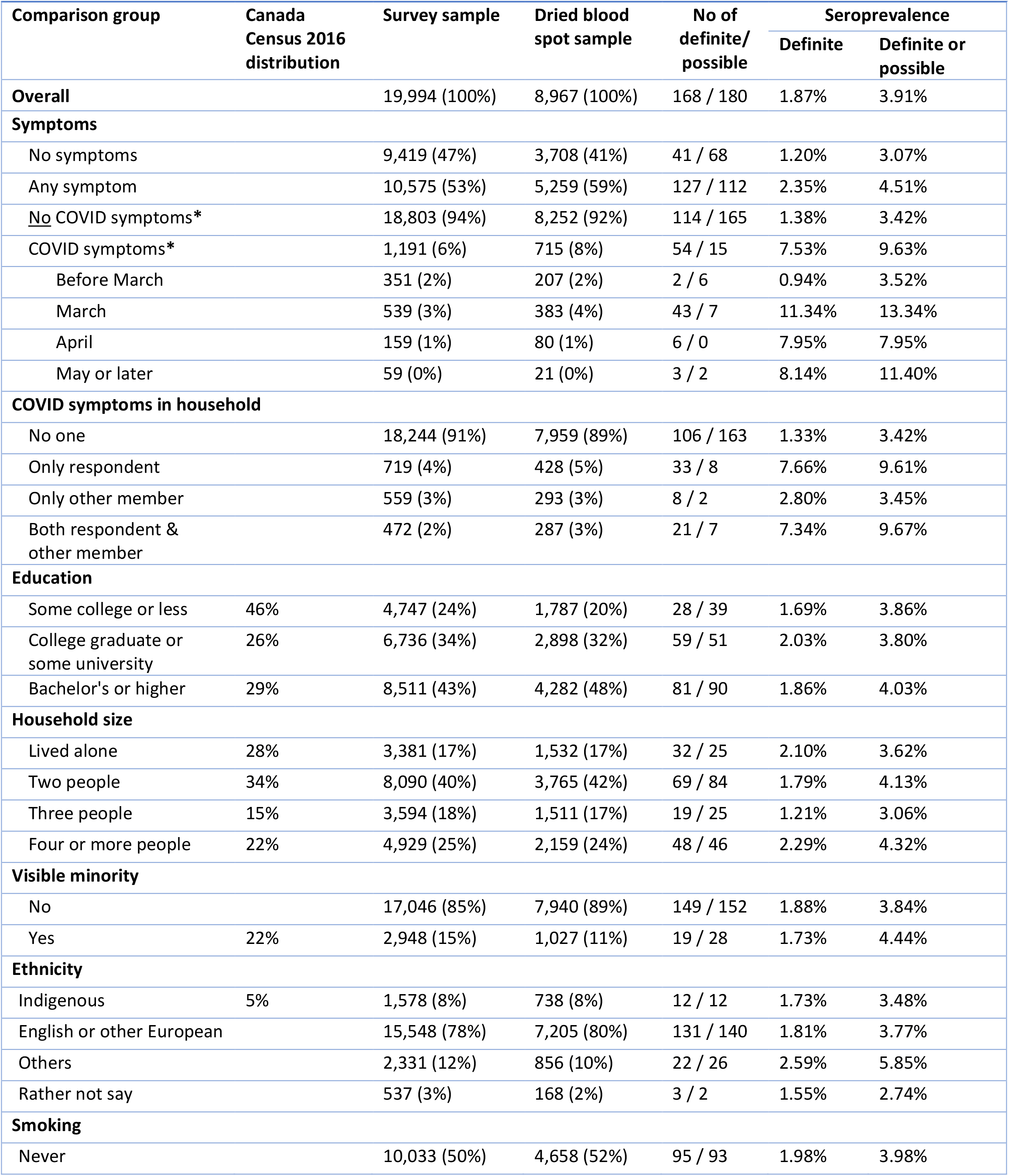
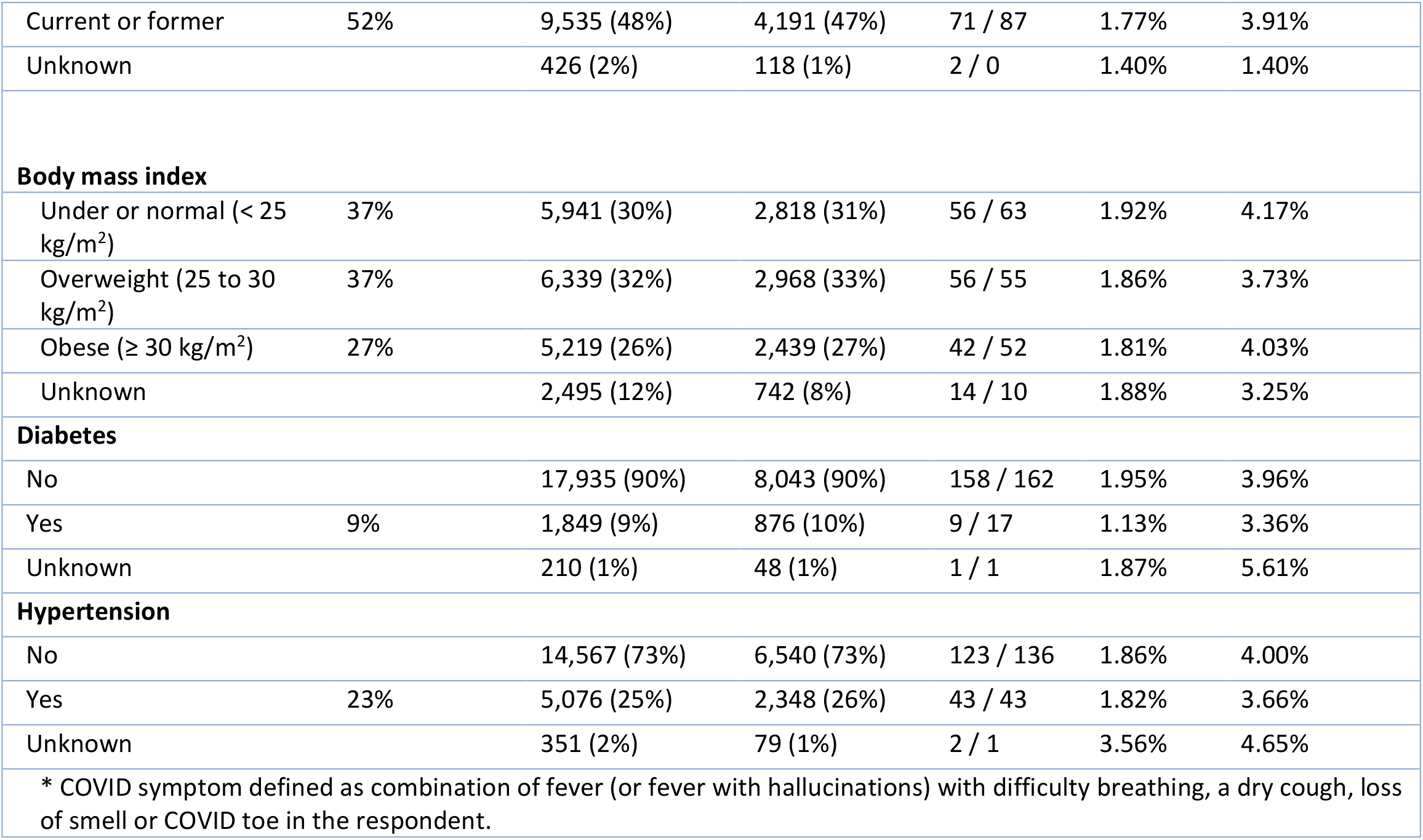
Representativeness of online poll, dried blood spot sample, and SARS-CoV-2 seroprevalence in Canada by individual traits, 2020

| Comparison group | Canada Census 2016 distribution | Survey sample | Dried blood spot sample | No of definite/ possible | Seroprevalence |  |
| --- | --- | --- | --- | --- | --- | --- |
|  |  |  |  |  | Definite | Definite or possible |
| **Overall** |  | 19,994 (100%) | 8,967 (100%) | 168 / 180 | 1.87% | 3.91% |
| **Symptoms** |  |  |  |  |  |  |
| No symptoms |  | 9,419 (47%) | 3,708 (41%) | 41 / 68 | 1.20% | 3.07% |
| Any symptom |  | 10,575 (53%) | 5,259 (59%) | 127 / 112 | 2.35% | 4.51% |
| <u>No</u> COVID symptoms* |  | 18,803 (94%) | 8,252 (92%) | 114 / 165 | 1.38% | 3.42% |
| COVID symptoms* |  | 1,191 (6%) | 715 (8%) | 54 / 15 | 7.53% | 9.63% |
| Before March |  | 351 (2%) | 207 (2%) | 2 / 6 | 0.94% | 3.52% |
| March |  | 539 (3%) | 383 (4%) | 43 / 7 | 11.34% | 13.34% |
| April |  | 159 (1%) | 80 (1%) | 6 / 0 | 7.95% | 7.95% |
| May or later |  | 59 (0%) | 21 (0%) | 3 / 2 | 8.14% | 11.40% |
| **COVID symptoms in household** |  |  |  |  |  |  |
| No one |  | 18,244 (91%) | 7,959 (89%) | 106 / 163 | 1.33% | 3.42% |
| Only respondent |  | 719 (4%) | 428 (5%) | 33 / 8 | 7.66% | 9.61% |
| Only other member |  | 559 (3%) | 293 (3%) | 8 / 2 | 2.80% | 3.45% |
| Both respondent & other member |  | 472 (2%) | 287 (3%) | 21 / 7 | 7.34% | 9.67% |
| **Education** |  |  |  |  |  |  |
| Some college or less | 46% | 4,747 (24%) | 1,787 (20%) | 28 / 39 | 1.69% | 3.86% |
| College graduate or some university | 26% | 6,736 (34%) | 2,898 (32%) | 59 / 51 | 2.03% | 3.80% |
| Bachelor's or higher | 29% | 8,511 (43%) | 4,282 (48%) | 81 / 90 | 1.86% | 4.03% |
| **Household size** |  |  |  |  |  |  |
| Lived alone | 28% | 3,381 (17%) | 1,532 (17%) | 32 / 25 | 2.10% | 3.62% |
| Two people | 34% | 8,090 (40%) | 3,765 (42%) | 69 / 84 | 1.79% | 4.13% |
| Three people | 15% | 3,594 (18%) | 1,511 (17%) | 19 / 25 | 1.21% | 3.06% |
| Four or more people | 22% | 4,929 (25%) | 2,159 (24%) | 48 / 46 | 2.29% | 4.32% |
| **Visible minority** |  |  |  |  |  |  |
| No |  | 17,046 (85%) | 7,940 (89%) | 149 / 152 | 1.88% | 3.84% |
| Yes | 22% | 2,948 (15%) | 1,027 (11%) | 19 / 28 | 1.73% | 4.44% |
| **Ethnicity** |  |  |  |  |  |  |
| Indigenous | 5% | 1,578 (8%) | 738 (8%) | 12 / 12 | 1.73% | 3.48% |
| English or other European |  | 15,548 (78%) | 7,205 (80%) | 131 / 140 | 1.81% | 3.77% |
| Others |  | 2,331 (12%) | 856 (10%) | 22 / 26 | 2.59% | 5.85% |
| Rather not say |  | 537 (3%) | 168 (2%) | 3 / 2 | 1.55% | 2.74% |
| **Smoking** |  |  |  |  |  |  |
| Never |  | 10,033 (50%) | 4,658 (52%) | 95 / 93 | 1.98% | 3.98% |
| Current or former | 52% | 9,535 (48%) | 4,191 (47%) | 71 / 87 | 1.77% | 3.91% |
| Unknown |  | 426 (2%) | 118 (1%) | 2 / 0 | 1.40% | 1.40% |
| **Body mass index** |  |  |  |  |  |  |
| Under or normal (< 25 kg/m<sup>2</sup>) | 37% | 5,941 (30%) | 2,818 (31%) | 56 / 63 | 1.92% | 4.17% |
| Overweight (25 to 30 kg/m<sup>2</sup>) | 37% | 6,339 (32%) | 2,968 (33%) | 56 / 55 | 1.86% | 3.73% |
| Obese (≥ 30 kg/m<sup>2</sup>) | 27% | 5,219 (26%) | 2,439 (27%) | 42 / 52 | 1.81% | 4.03% |
| Unknown |  | 2,495 (12%) | 742 (8%) | 14 / 10 | 1.88% | 3.25% |
| **Diabetes** |  |  |  |  |  |  |
| No |  | 17,935 (90%) | 8,043 (90%) | 158 / 162 | 1.95% | 3.96% |
| Yes | 9% | 1,849 (9%) | 876 (10%) | 9 / 17 | 1.13% | 3.36% |
| Unknown |  | 210 (1%) | 48 (1%) | 1 / 1 | 1.87% | 5.61% |
| **Hypertension** |  |  |  |  |  |  |
| No |  | 14,567 (73%) | 6,540 (73%) | 123 / 136 | 1.86% | 4.00% |
| Yes | 23% | 5,076 (25%) | 2,348 (26%) | 43 / 43 | 1.82% | 3.66% |
| Unknown |  | 351 (2%) | 79 (1%) | 2 / 1 | 3.56% | 4.65% |
| * COVID symptom defined as combination of fever (or fever with hallucinations) with difficulty breathing, a dry cough, loss of smell or COVID toe in the respondent. |  |  |  |  |  |  |

In the entire polling sample, 10,575 respondents experienced at least one of the survey symptoms, and 1,191 (6% of the entire cohort) met the study definition of COVID symptom positivity. The predictors of COVID symptom-positivity were broadly similar to the predictors of seropositivity. In multivariate analyses comparing definite seropositives to negatives among those who experienced any symptom (COVID or otherwise) since February, seroprevalence was higher in those with COVID symptoms, in those who reported symptoms in March, and in those who lived alone (Figure 1). Seroprevalence was lower for age ≥70, and lower in provinces other than Ontario. Adjusted for other variables, COVID symptoms were more common in women than men, in visible minorities or in Indigenous populations than in other ethnic groups, in current or former smokers than non-smokers, and among self-reported hypertensives and diabetics (Appendix Tables 2-3).

**Figure 1.**
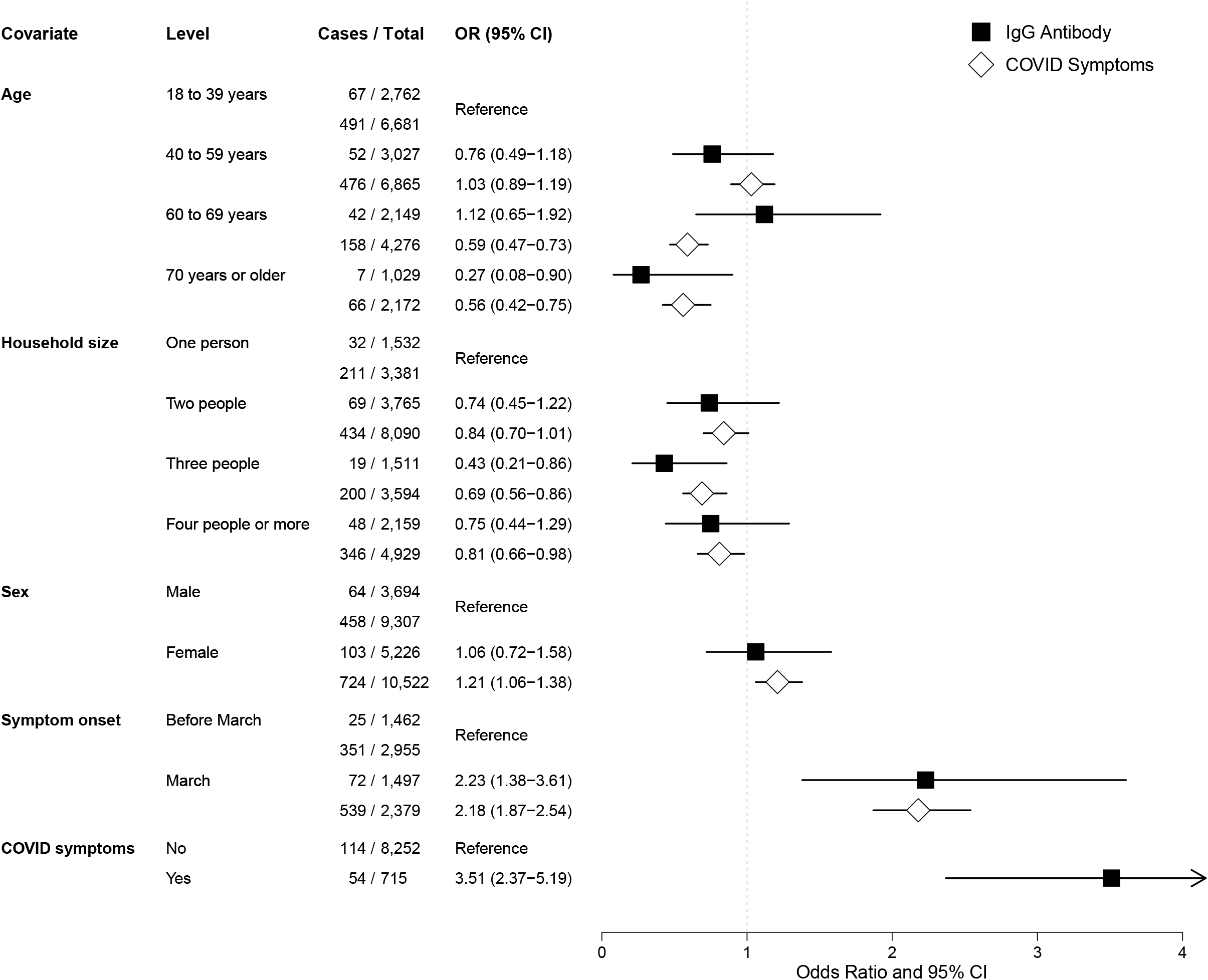
Determinants of IgG antibody (definite positives) and COVID symptoms in the Ab-C study in Canada.

## Discussion

Overall SARS-CoV-2 seroprevalence among Canadian adults in the community was low during the first viral wave, consistent with modest COVID mortality outside of nursing homes, reflecting some success in containing the community spread of infection. Adult seroprevalence nationally was 1.7-3.5%, with Ontario, the most populous province, having the highest seroprevalence. This suggests that roughly 0.54-1.08 million adult Canadians were infected, nearly half of them young adults. The Canadian adult seroprevalence was lower than those reported among adults in national seroprevalence studies in England and Spain, ^21-23^ and from convenience sampling in the US,^24^ but higher than that in a national study in Iceland.^25^ The overall and age-specific IFRs were lower than estimated in multi-country reviews.^11,26^ As of the end of August, about 150,000 Canadians were PCR-confirmed cases, suggesting a crude ratio of 4 to 8 infected to each confirmed PCR-positive cases, which was a lower ratio than those at comparable time periods in England, Spain,^21-23^ or the US.^24^

Despite reasonable control of community transmission during the first viral wave, Canada had a large excess of nursing home deaths. Based on the number of deaths, we estimate, crudely, that about 13-17% of nursing home residents became infected, and that the nursing home death rate was more than 70 times higher than that in comparably-aged adults living outside of these facilities, compatible with other Canadian and international comparisons.^27^

Asymptomatic persons constituted about a quarter of definite seropositives, but the proportion was much higher among the elderly. We did not directly measure seroprevalence in nursing home settings, and in addition to there possibly being more asymptomatic transmission in the elderly, the transmission dynamics vary greatly across facilities. Nursing homes remain vulnerable in the current viral wave, accounting for two-thirds of the approximately 9,000 COVID deaths from September 1, 2020 to February 1, 2021.^1,10^

This study documents a low seroprevalence of IgG antibodies in Canada, consistent with earlier convenience samples from blood donors and residual sera from public health laboratory specimens,^28-31^ suggesting that in the absence of other evidence about widespread cellular immunity,^31^ nearly all adult Canadians will need to be vaccinated. The Ab-C study did not collect data on children.

Despite the relatively low IFRs observed in Canada, our study helps to counter earlier ill-conceived notions that argued for “natural herd immunity” by permitting adults outside of nursing home residents to become infected.^33^ Assuming 60% of each age group shown in Table 1 would need to be infected, and multiplying this by IFRs in the same table leads to an estimate of between 40,000 and 139,000 adult deaths outside of nursing home residents. Moreover, it is nearly impossible to shield nursing home residents from community-based transmission, as has been clear in the current viral wave.

As vaccination programs will need to achieve high population coverage, seroprevalence should rise and continued antibody testing (using differences in antibody responses that can distinguish natural from vaccine-induced immunity) is needed to assess the impact of vaccines, including on different sub-populations. Our results demonstrate that the Ab-C home-based DBS collection is highly practicable and affordable, and compatible with physical distancing requirements for COVID control.

The Ab-C study achieved reasonable representativeness of the Canadian adult population, and our cohort was comparable in the prevalences of obesity, smoking, and other risk factors for COVID. We used a highly sensitive ELISA assay that has previously been shown to measure IgG antibodies for several months after infection.^7^ We attempted to minimize false positives by applying stringent cut-off levels to define high specificity. Moreover, we collected DBS samples sufficiently early (June-August) to provide a reliable, comprehensive assessment of Canada’s experience with the first viral wave (which peaked in terms of COVID symptoms in March; PCR-confirmed cases in April and deaths in May^1^; Appendix Figure 10).

However, the study has some limitations. First, while our assay results were likely to be consistent throughout the Ab-C study population, the results are less comparable to seroprevalence studies using other assays. Ideally, testing with multiple assays should be done to reduce false positives in particular at low prevalence. However, since nearly all SARS-CoV-2 assays are recent, with less than a year for development and deployment, cross-comparisons that allow multiple assays are challenging.^35^ To improve comparisons over time and across assays, we recommend that national authorities organize a testing scheme with defined, blinded sample panels that can be provided to relevant laboratories. A similar strategy was successful at improving HIV diagnostics.^36^ Second, we could not directly measure seropositivity in residents in nursing homes, who accounted for most COVID deaths in Canada. By necessity, we used IFRs from French nursing homes, which were broadly similar in their demographics.^27^ The mortality-based and direct seroprevalence studies may not be comparable. Third, we may have overestimated the proportion of respondents who were asymptomatic, particularly at older ages, as we elicited a limited range of symptoms. The asymptomatic proportion of a quarter among definite seropositives was similar to that reported in England,^23^ higher than in a systematic review,^37^ and consistent with another that found more asymptomatic infections at older ages.^38^ Finally, our sample enrolled fewer lower education groups than planned. We adjusted for differences in education, but there might well be unrecorded variables related to COVID risk that affected participation or willingness to provide a DBS.

The Ab-C study provides a useful benchmark for Canada to identify infection levels from the first COVID wave. Repeat assessments in the same cohort have begun in the first quarter of 2021, and will be repeated in the third quarter of 2021. These assessments will enable documentation of incidence of infection during the intervening months and monitor cumulative seroprevalence and IFRs in the ongoing second viral wave and the impact of vaccines.

## Data Availability

Data available upon request and subject to approval from Unity Health Toronto Ethics Review Board and the Canadian COVID-19 Immunity Task Force.

## Acknowledgements

We thank the thousands of Canadians who participated in the Ab-C study. Funding was provided by an unrestricted grant from Pfizer Global Medical Grants and from Unity Health Toronto and the Canadian COVID-19 Immunity Task Force. Funding for the development of the assays in the Gingras lab was provided by Royal Bank of Canada, QuestCap, and the Krembil Foundation. The robotics equipment at the Network Biology Collaborative Centre at the Lunenfeld-Tanenbaum Research Institute is supported by the Canada Foundation for Innovation, the Ontario Government, Genome Canada, and Ontario Genomics (OGI-139). EuroImmun Diagnostics (Sean McFadden) supported the testing platform at St. Josephs Health Centre/Unity Health. Statistics Canada (Ron Gravel, Scott McLeish) provided analyses on deaths by location. The National Research Council Canada (Yves Durocher) supplied ELISA reagents, and the National Microbiology Laboratory of Canada provided samples for DBS testing.

## Authors

The members of the writing and implementation committees are as follows: Xuyang Tang, Abha Sharma, Maria Pasic, Patrick Brown, Karen Colwill, Chaim Birnboim, Nico Nagelkerke, Isaac Bogoch, Craig Schultz, Leslie Newcombe, Justin Slater, Peter S Rodriguez, Guowen Huang, Sze Hang Fu, Catherine Meh, Chee Nee Wu, Rupert Kaul, Marc-André Langlois, Ed Morawski, Andy Hollander, Demetre Eliopoulos, Benjamin Aloi, Teresa Lam, Kento T Abe, Lauren Caldwell, Miriam Barrios-Rodiles, Mahya Fazel-Zarandi, Ronald Weingust, Jenny Wang, Bhavisha Rathod, Divya Raman Santhanam, Eo Rin Cho, Kathleen Qu, Shreya Jha, Vedika Jha, Wilson Suraweera, Varsha Malhotra, Richard Wen, Samir Sinha, Angus Reid, Anne-Claude Gingras, Pranesh Chakraborty, Arthur S Slutsky, Prabhat Jha.

## Author Affiliations

From: the Centre for Global Health Research, Unity Health Toronto and University of Toronto (X Tang PhD, A Sharma MPH, P Brown, PhD, C Birnboim, MD, N Nagelkerke, PhD, C Schultz MSc, L Newcombe BSc, J Slater MSc, PS Rodriguez MSA, G Huang PhD, SH Fu MSA, C Meh MPH, CN Wu MSc, DR Santhanam BSc, ER Cho PhD, K Qu MPH, S Jha BMus, V Jha, W Suraweera MSc, V Malhotra PhD, R Wen MSc, Prof P Jha); Sinai Health Toronto (K Colwill PhD, KT Abe MS, L Caldwell BSc, M Barrios-Rodiles PhD, M Fazel-Zarandi, J Wang MSc, B Rathod BSc, Prof S Sinha MD, Prof AC Gingras PhD); St. Joseph’s Health Centre, Unity Health, Toronto (M Pasic PhD, R Weingust MSc); University Health Network (I Bogoch MD, Prof R Kaul MD); University of Ottawa (Prof P Chakraborty MD, Prof MA Langlois PhD), Angus Reid Institute (E Morawski MA, A Hollander BA, D Eliopoulos BA, B Aloi MPP, T Lam PhD, A Reid PhD); Unity Health Toronto (Prof AS Slutsky MD).

## Appendix

### Section 1: Study Flow, Sampling, and High-Burden Regions

**Appendix Figure 1:**
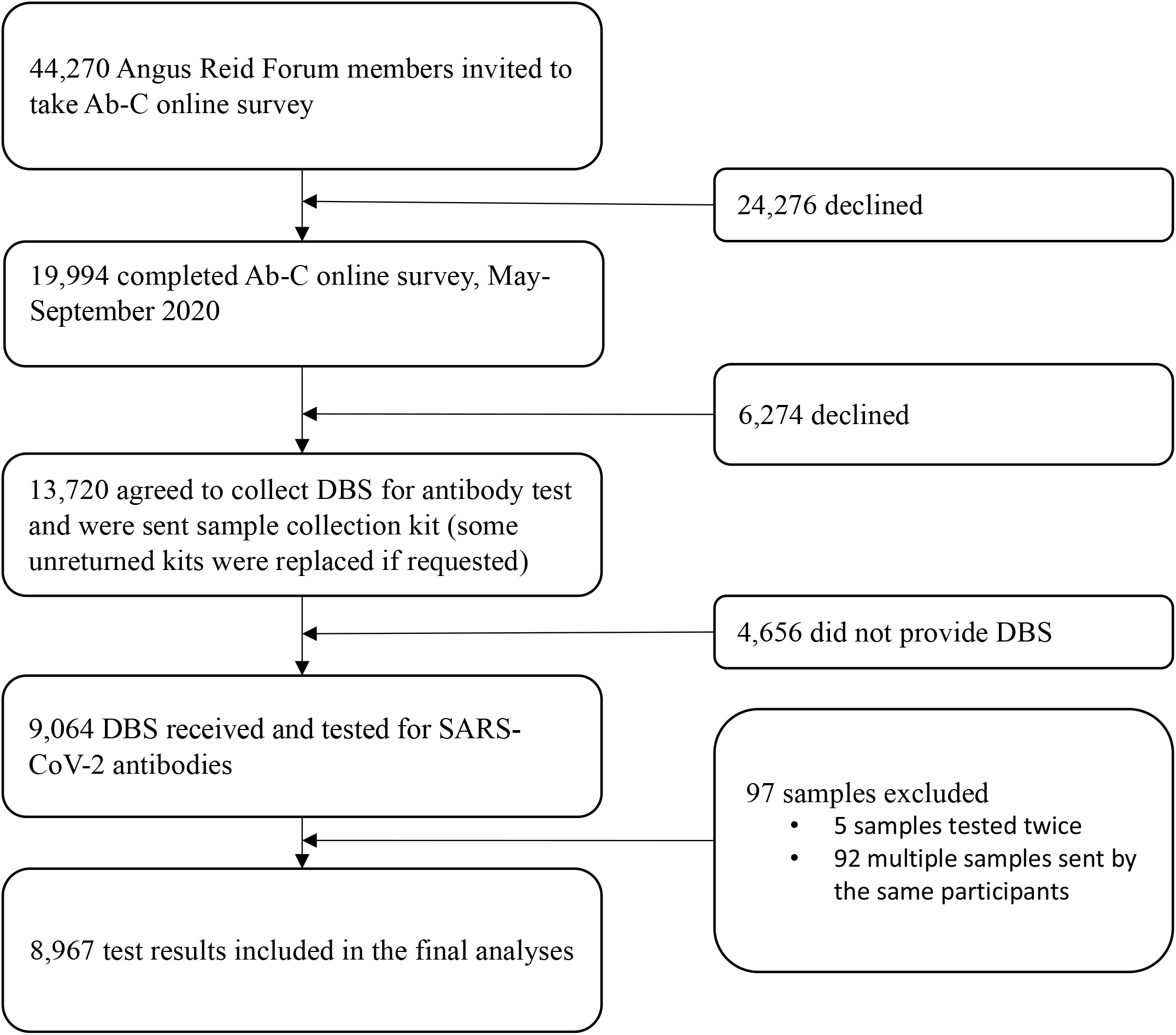
Study flow of the overall sampling.

#### Angus Reid Forum Sampling

The sample for the Ab-C study was drawn from the Angus Reid Forum (ARF) online Canada-wide panel operated by the Angus Reid Group. ARF employs a multi-stage stratified sample that begins with 300 regional sample points roughly analogous to Canada’s federal political riding boundaries. The sample framework is further stratified by gender, age, and education within composite regions. All national studies are conducted in Canada’s two official languages, English and French. ARF studies typically achieve response rates of 35-45%. The Ab-C response rate was 49%.

The sampling design was to first stratify by census metropolitan area, age groups (18-34, 35-54, 55+), sex (male, female), and education (high school education or lower, some college or college or technical degree, some university, or university degree). Panel members who met the stratification criteria were invited to complete the survey.

#### High-Burden Regions

Given the concerns about rising COVID cases during a second wave, we conducted a spatial analysis of PCR-confirmed COVID cases in the 93 public health units in Canada, using data up to July 2020. A Poisson spatial regression had case counts as the response variable, an age-sex adjusted expected count as an “offset,” and predictor variables of lung cancer incidence (as a proxy for smoking exposure, a risk factor for severe COVID), unemployment, and proportion of visible minorities. A spatial random effect term, where each unit has conditional dependence with each of its neighbors, was added to account for spatial clustering. This analysis identified 17 health units at notably higher risk of COVID incidence (Appendix Figure 2), which had 28.5% visible minorities, twice the 14.5% rate of visible minorities for the whole of Canada.

**Appendix Figure 2.**
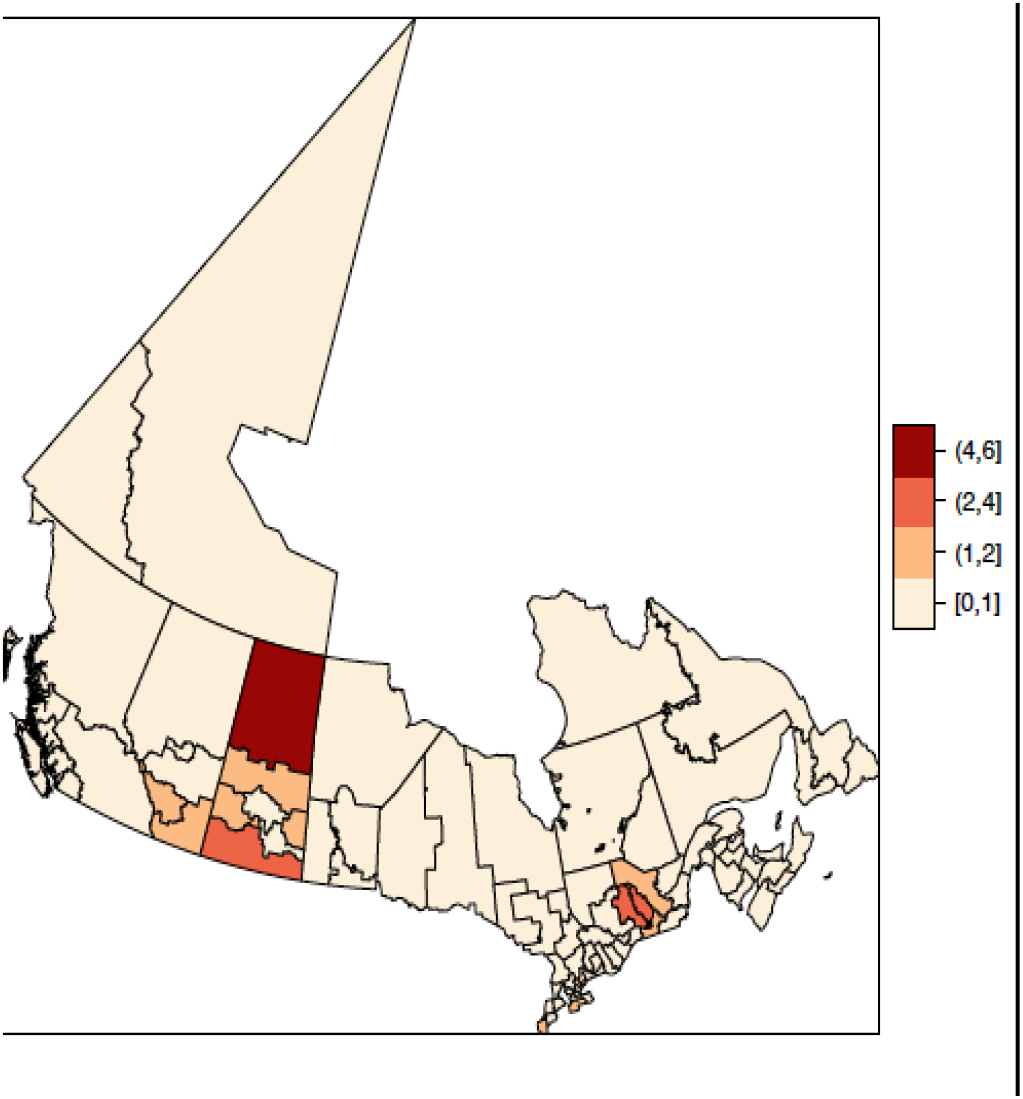
Map of 17 high-burden COVID areas based on regression analyses of PCR testing rates. Right scale represents the relative risk of COVID cases

### Section 2: Laboratory Testing Procedures

For DBS controls, a 6 mm punch (28.26 mm^2^) was taken from one DBS. The antibodies were eluted from the punch in 125 ⍰l of PBS + 0.1% Tween (PBS-T) and 1% Triton X-100 for a final area to volume ratio of 0.226 mm^2^/⍰l. To maintain the equivalent ratio with the 4.7 mm punches (17.3 mm^2^) used for Ab-C participants, the antibodies were eluted in 77 ⍰l PBS-T with 1% Triton X-100. Punches were incubated in elution buffer for a minimum of 4 hours with gentle shaking (150 RPM). The samples were then centrifuged at 1000 g for 30 seconds.

Automated chemiluminescent ELISA assays were performed as previously described on a ThermoFisher Scientific F7 robotic platform^1^ with a few modifications. Briefly, LUMITRAC 600 high-binding white polystyrene 384-well microplates (Greiner Bio-One #781074, VWR #82051-268) were pre-coated overnight with 10 ⍰l/well of spike trimer (50 ng SmT1, National Research Council Canada (NRC)). After washing (all washes were 4 times with 100 ⍰l PBS-T), wells were blocked for 1 hour in 80 ⍰l 5% Blocker BLOTTO (Thermo Fisher Scientific, #37530) and then washed. 10 ⍰l of sample (2.5 ⍰l of DBS eluate diluted in 1% final Blocker BLOTTO in PBS-T, which represents a 1:50 dilution of eluate from a 6 mm punch) was added to each well and incubated for 2 hours at room temperature. After washing, 10 ⍰l of a human anti-IgG fused to HRP (Ig#5, supplied by NRC, final of 0.9 ng/well) diluted in 1% final Blocker BLOTTO in PBS-T was added to each well followed by a 1-hour incubation at room temperature. After 4 washes, 10 ⍰l of SuperSignal ELISA pico chemiluminescent substrate (diluted 1:4 in water) was added to each well and incubated for 5 min at room temperature. Luminescence was read on an EnVision (PerkinElmer) plate reader at 100 ms/well using an ultra-sensitive detector.

Each 384-well assay plate included replicates of a standard dilution curve of VHH72 (a human anti-spike IgG antibody^2^), positive and negative master mixes of pooled serum samples, human IgG negative control (Sigma, I4506), and blanks as controls. Samples were processed over 6 separate runs. Negative and positive DBS controls were included in 4 of the runs.

Blank values were subtracted from all raw reads (counts per second of luminescence), and the values were expressed as a ratio of the 0.0156 g/ml point of the standard curve of VHH72. The stricter threshold for positives was determined by calculating the mean plus three standard deviations of a series of negative control DBS samples. The lenient threshold for possible samples was calculated using the presumed negative distribution of the Ab-C samples. Samples with a relative ratio of > 0.05 and < 0.3 were selected, and the mean plus three standard deviations of these samples was used for the cut-off. Data were analyzed and plots were generated in R using version 4.0.1^3^.

Appendix Figure 3 shows the reproducibility of the standard dilution curves of VHH72 (in μg/ml) across the different runs (run 1 to run 6). The reproducibility in signal (blank-adjusted counts per second) between runs in the linear range and in the more dilute concentrations is high. More variation in signal is observed for the less dilute concentrations outside of the linear range.

**Appendix Figure 3.**
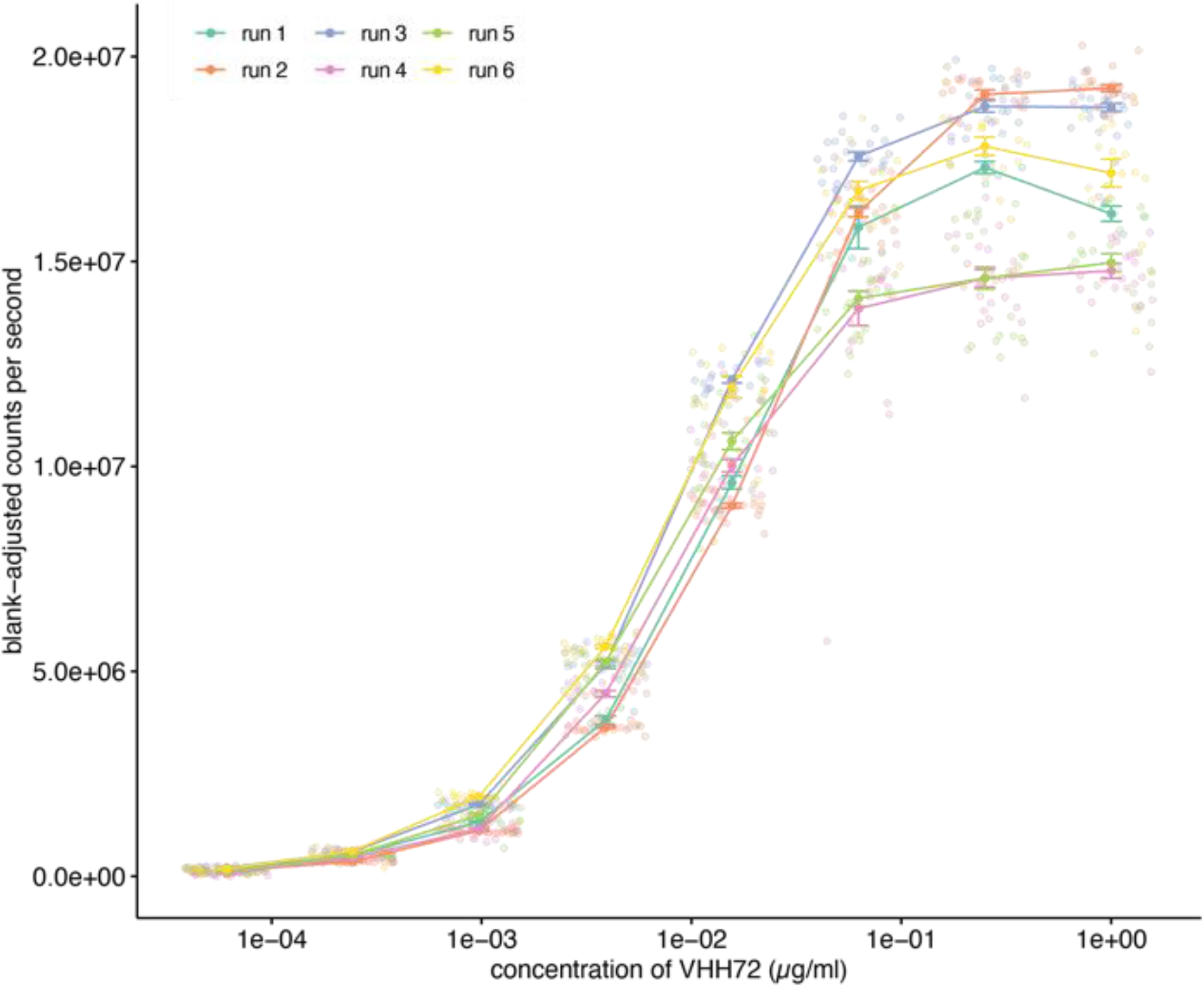
Reproducibility of the standard dilution curves of the anti-spike antibody VHH72 (in ⍰g/ml)

Appendix Figure 4 shows the normalized values of the mean of controls included in each assay run. The reproducibility among batches is high between runs at points along the standard curve (samples 0.0000609 to 1 µg/ml) up to the midpoint to which values were normalized (0.0156 µg/ml). This is important because the calls for positive/negative fall within this range. In addition, two dilutions of pooled serum samples from patients negative and positive for anti-spike IgG were run.

**Appendix Figure 4.**
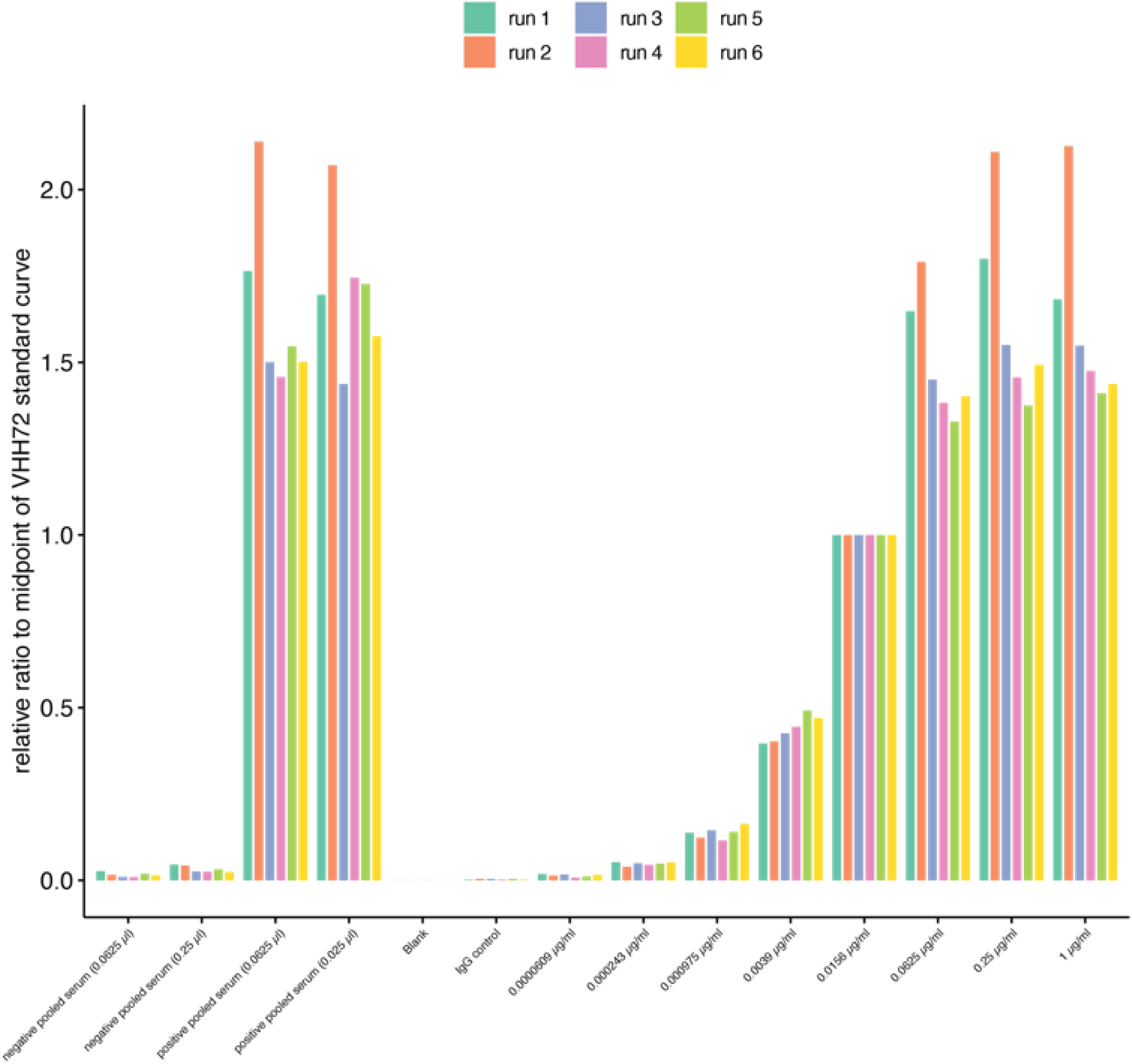
Normalized values of the mean of controls to the 0.0156 µg/ml of the standard curve (anti-spike antibody VHH72)

Appendix Figure 5 shows the reproducibility between technical replicates in the ELISA assay. Run 1 was processed in technical duplicates. Blank-adjusted chemiluminescent values are expressed as a relative ratio to the 0.0156 µg/ml midpoint of the VHH72 standard curve (n = 304). Spearman correlation analysis was performed, and a coefficient of 0.84 indicates that the replicates are very strongly correlated (*p* < 0.0001). The shaded area indicates the 95% confidence interval.

**Appendix Figure 5.**
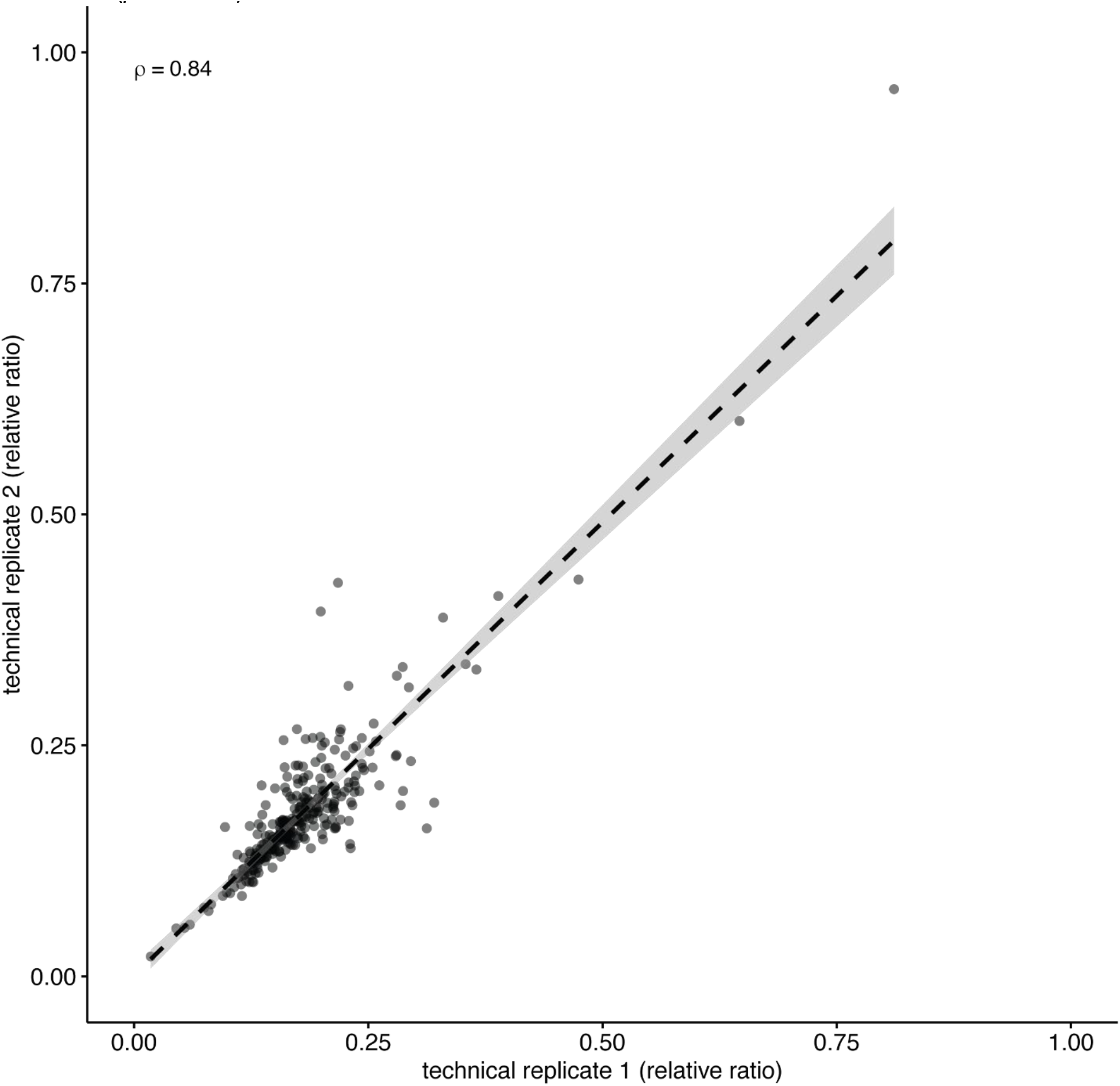
Reproducibility between duplicates in the automated ELISA.

Appendix Figure 6 shows a density plot of the normalized Ab-C samples, and the rug plot along the bottom shows each data point along the x-axis. The large peak represents samples with low signal and an extended tail marks samples of increased signal. The green line represents the 0.3899 cut-off value, which is 3 SD from the mean of the known negative DBS samples. The yellow line is the mean plus 3 SD from the presumed negatives of the Ab-C dataset with a cut-off value of 0.2724. The dotted line indicates the median relative ratio value of the entire dataset.

**Appendix Figure 6.**
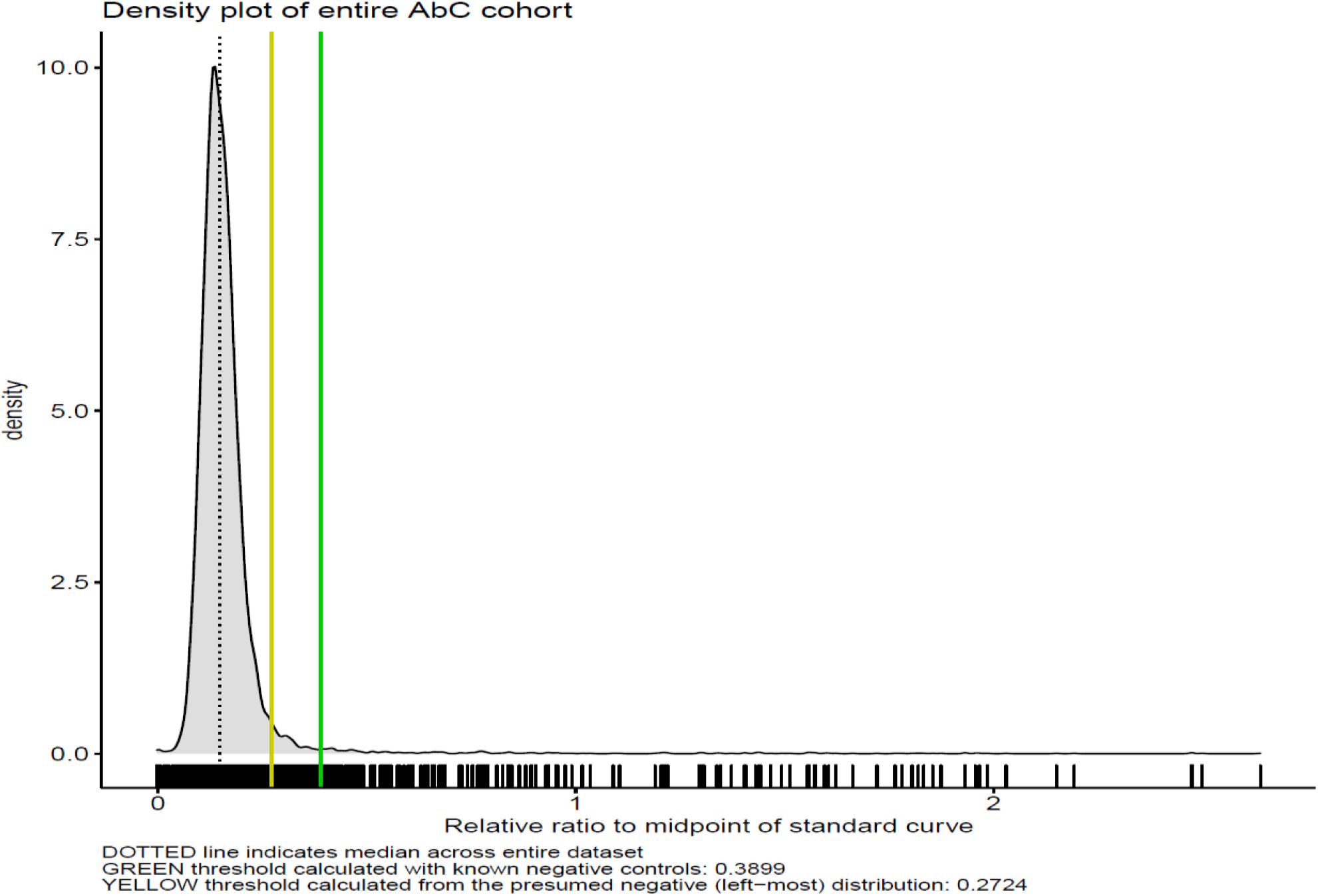
Density of the normalized Ab-C samples.

Appendix Figure 7 ranks all the Ab-C samples from the strongest to the weakest signal. The green line denotes the threshold calculated from known negative DBS samples and the yellow line denotes the threshold calculated from the presumed negative distribution. Samples are color-coded based on their call. Applying these thresholds to the Ab-C DBS sample, we have 169 positives (1.9%), 182 possible (2%), and 8713 negative (96%) readings. The inset graph is a zoomed in version of samples close to the two-cut offs.

**Appendix Figure 7.**
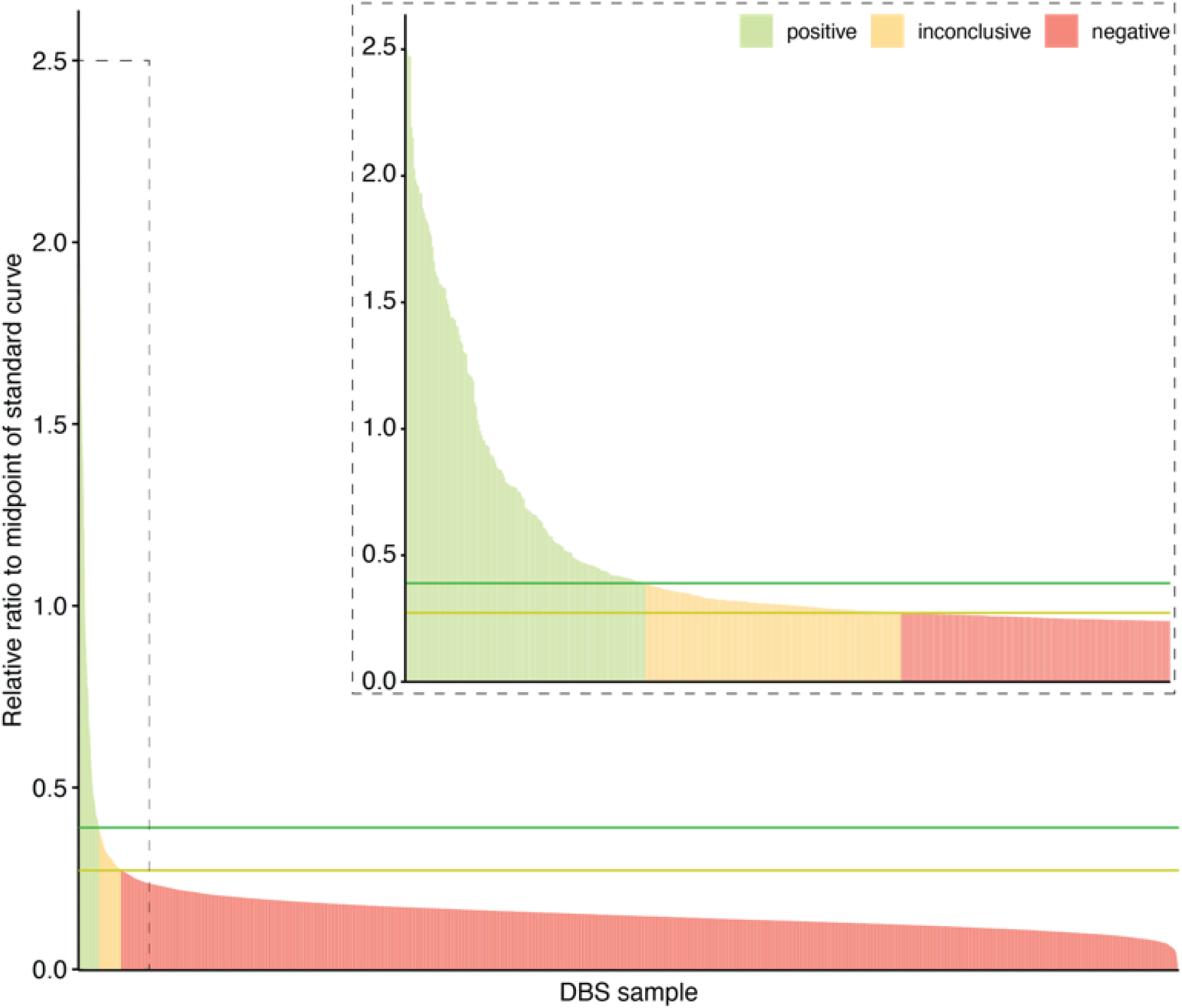
Ranking of All Ab-C Samples from the Strongest to the Weakest Signal.

### Section 3: Analyses of SARS-CoV-2 Infection and COVID Symptom Positivity

#### Measures and variables for logistic regression

The variables examined included sex, COVID symptom positivity (defined below), age group (18-39, 40-59, 60-69, and 70+), and province (Ontario, Quebec, British Columbia and Yukon, Atlantic Canada, and Prairie Canada and Northwest Territories).

#### COVID Symptom Positivity

Several retrospective studies that enrolled more than 100 COVID patients in hospitals in mainland China early in the pandemic have identified fever^4–10^ and cough (dry or unspecified)^4,6–10^ as commonly associated with the infection. Other symptoms experienced by patients in China include a lack of appetite,^5^ general weakness,^5^ malaise,^9^ myalgia or fatigue,^6,9,10^ diarrhea,^5^ excess sputum production,^6^ shortness of breath,^7,9^ and sore throat.^10^ A study that examined 1,420 COVID patients from 18 hospitals in Europe found that headache and loss of smell were present in over 70% of the cases.^11^ A cutaneous manifestation appearing similar to skin rash (“COVID toe”) was common among 375 COVID patients in Spain.^12^ The online Ab-C survey questions about COVID symptoms were developed based on these findings.

We asked respondents if they had experienced any of the following symptoms that were not related to a condition or illness that they dealt with chronically: difficulty breathing, fever, mild dry cough, severe dry cough (“keeps you from sleeping”), sore throat, frequent sneezing, loss of sense of smell or taste, fever with hallucinations, unusual or disturbed sleep, loss of appetite, dizziness, and red, purple, pink toes with swelling. For each symptom, the possible responses were “Yes, had this but it went away,” “Yes, I still have this,” and “No, have not had this.” We also asked respondents for the month they first had these symptoms, and if they were still having any of them. We asked the respondents the same questions about their household members’ experience with the same symptoms. We asked if they worked in a list of high-exposure occupations (e.g., health care staff or transit workers) and if they had visited health care or nursing home facilities.

We created separate COVID symptom positivity variables for respondents and for household members. We defined COVID symptom-positive as a combination of fever (or fever with hallucinations) with difficulty breathing, a dry cough, loss of smell, or “COVID toe.” We created a categorical variable to indicate if no one in the household, only the respondent, only other household member(s), or both the respondent and household member(s) were COVID symptom-positive.

#### Demographics and health

Respondents reported their birth year and month, sex (male/female/prefer to self-describe), highest level of education (some elementary or high school/high school graduate/some college or trade school/graduated from college or trade school/some university/university undergraduate degree/university graduate degree), height, weight, current smoking status (never smoked/smoked daily or occasionally/used to smoke but quit/don’t know/prefer not to answer), diagnosis of diabetes (yes/no/don’t know/prefer not to answer), and diagnosis of high blood pressure (yes/no/don’t know/prefer not to answer). We calculated body mass index (BMI) from self-reported height and weight, and categorized BMI as normal or underweight (BMI < 25 kg/m^2^), overweight (25 kg/m^2^ ≤ BMI < 30 kg/m^2^), or obese (BMI ≥ 30 kg/m^2^).^13^ The Angus Reid Forum membership database has existing information on the respondents’ age in years, ethnicity (including whether they self-identify as a visible minority), and region of residence.

##### Statistical analysis

We defined seropositive cases as definite using a cut-off of more than 3 standard deviations from control values. The logistic regression analyses defined these as cases and compared only the definite positives to the uninfected controls. We also present the proportion of respondents who were COVID symptom-positive by month of onset and age group, and the distribution of COVID symptom positivity in households by month of onset and age group. We used simple logits to calculate the likelihood of COVID symptom positivity by month in the overall sample, adjusting for known demographic and health risk factors. We excluded responses from 638 respondents with questionable self-reported height (< 100 or > 221 centimeters) and 1,080 respondents with questionable self-reported weight (< 30 or > 200 kilograms) from the logit but not for other analyses that do not require weight and height information. We used Stata SE 13^14^ to conduct our analyses.

##### Predictors of SARS-CoV-2 seropositivity

Appendix Table 1 provides the odds ratios for the predictors of the risk of seropositivity. The main predictors of increased odds of seropositivity were COVID symptoms (OR = 3.51; 95% CI 2.37-5.19) and any symptom onset in March (OR = 2.23, 1.38-3.61). The main predictors of reduced odds of seropositivity were provinces other than Ontario (all OR <0.46), 70 years or older (OR = 0.27, 0.08-0.90), and being in a household with three people (OR = 0.43, 0.21-0.86).

**Appendix Table 1.**
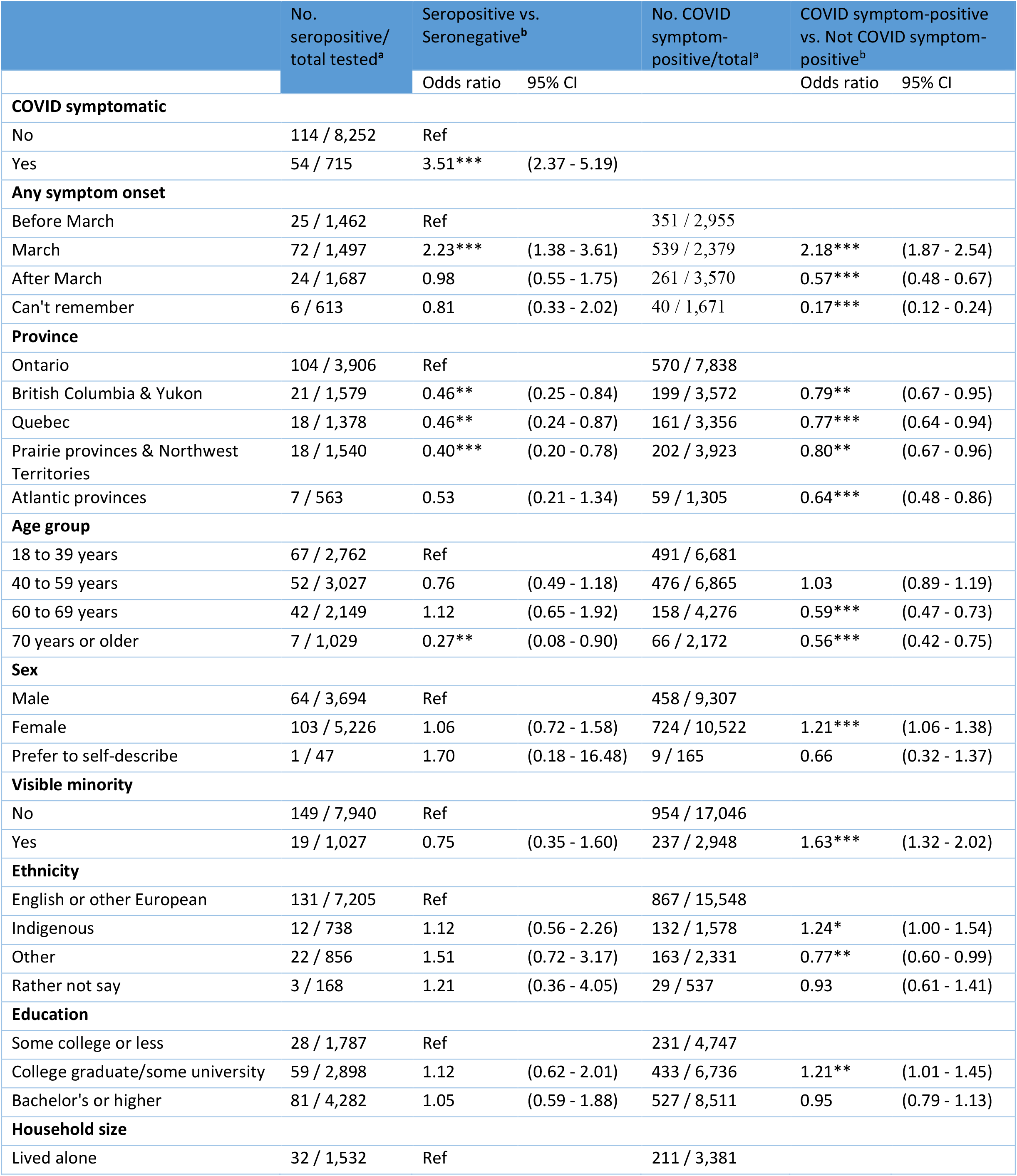
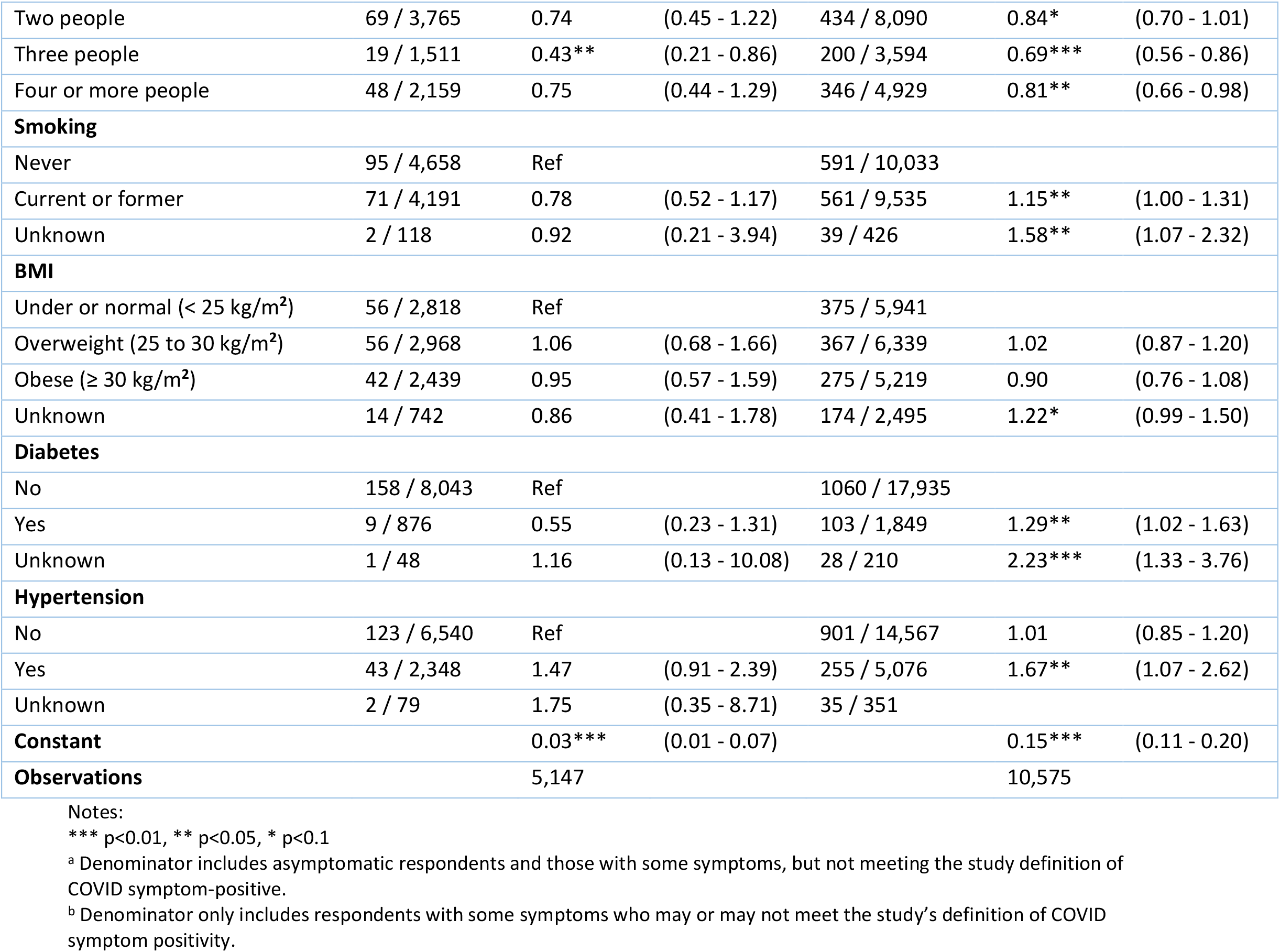
Respondents’ odds ratios of seropositivity for selected variables

##### Predictors of asymptomatic seropositivity

Appendix Table 2 provides the odds ratios for having asymptomatic (meaning no symptoms at all) seropositivity (24% asymptomatic among definite seropositives and 31% asymptomatic among definite and possible seropositives). As the number of asymptomatic respondents was small using the definite seropositives, results are shown for both definite and definite and possible seropositives. After adjusting for age, sex, ethnicity, education, province of residence, BMI, smoking status, and history of diabetes and hypertension, respondents living in the Prairie provinces and the Northwest Territories had higher odds of reporting no symptoms (OR = 4.85, 1.03-22.84 among definite seropositives and OR = 2.58, 1.08-6.13 among definite and possible seropositives) than the reference group of respondents living in Ontario. Respondents in British Columbia and Yukon had higher odds of having no symptoms (OR = 2.30, 1.02-5.19) than Ontarian respondents among definite and possible seropositives. Those aged 70+ had higher odds of reporting COVID symptoms (OR = 10.39, 0.86-125.90 among definite positives and OR = 5.39, 1.55-18.69 among definite and possible seropositives) compared to respondents aged between 18 and 39.

**Appendix Table 2.**
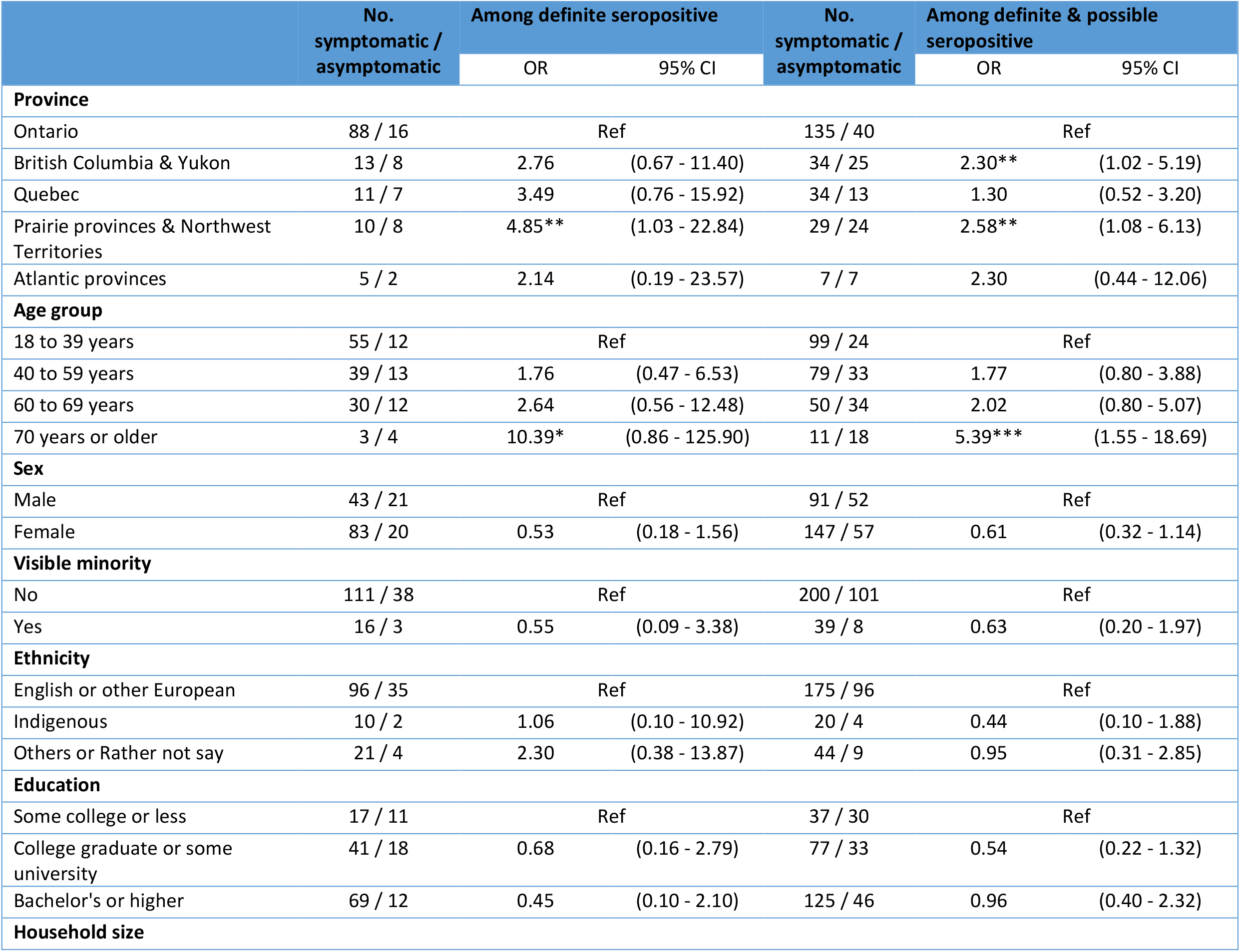
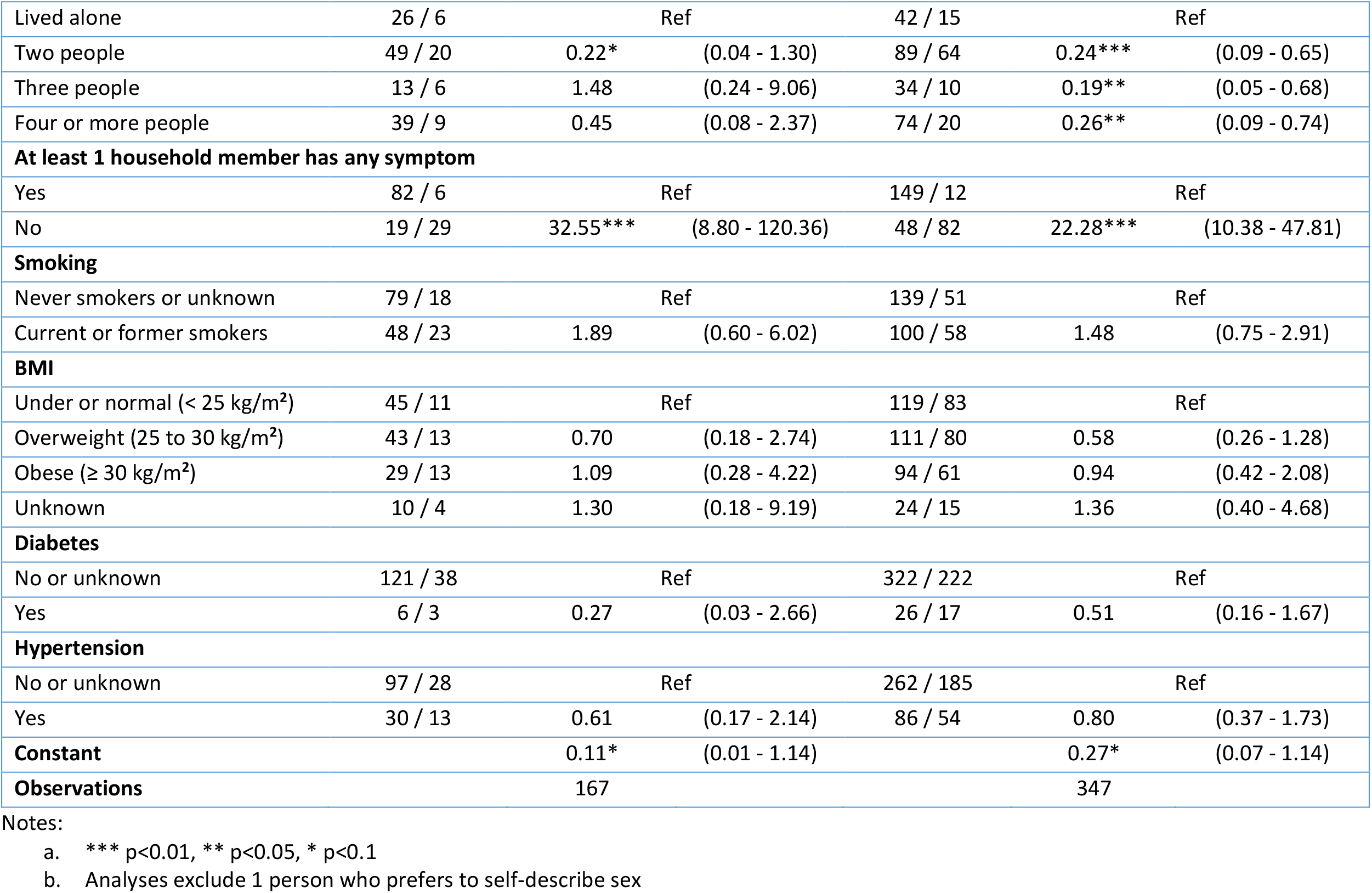
Respondents’ odds ratios of asymptomatic seropositivity for selected variables

|  | No.<br>symptomatic /<br>asymptomatic | Among definite seropositive |  | No.<br>symptomatic /<br>asymptomatic | Among definite & possible<br>seropositive |  |
| --- | --- | --- | --- | --- | --- | --- |
|  |  | OR | 95% CI |  | OR | 95% CI |
| Province |  |  |  |  |  |  |
| Ontario | 88 / 16 |  | Ref | 135 / 40 |  | Ref |
| British Columbia & Yukon | 13 / 8 | 2.76 | (0.67 - 11.40) | 34 / 25 | 2.30** | (1.02 - 5.19) |
| Quebec | 11 / 7 | 3.49 | (0.76 - 15.92) | 34 / 13 | 1.30 | (0.52 - 3.20) |
| Prairie provinces & Northwest Territories | 10 / 8 | 4.85** | (1.03 - 22.84) | 29 / 24 | 2.58** | (1.08 - 6.13) |
| Atlantic provinces | 5 / 2 | 2.14 | (0.19 - 23.57) | 7 / 7 | 2.30 | (0.44 - 12.06) |
| Age group |  |  |  |  |  |  |
| 18 to 39 years | 55 / 12 |  | Ref | 99 / 24 |  | Ref |
| 40 to 59 years | 39 / 13 | 1.76 | (0.47 - 6.53) | 79 / 33 | 1.77 | (0.80 - 3.88) |
| 60 to 69 years | 30 / 12 | 2.64 | (0.56 - 12.48) | 50 / 34 | 2.02 | (0.80 - 5.07) |
| 70 years or older | 3 / 4 | 10.39* | (0.86 - 125.90) | 11 / 18 | 5.39*** | (1.55 - 18.69) |
| Sex |  |  |  |  |  |  |
| Male | 43 / 21 |  | Ref | 91 / 52 |  | Ref |
| Female | 83 / 20 | 0.53 | (0.18 - 1.56) | 147 / 57 | 0.61 | (0.32 - 1.14) |
| Visible minority |  |  |  |  |  |  |
| No | 111 / 38 |  | Ref | 200 / 101 |  | Ref |
| Yes | 16 / 3 | 0.55 | (0.09 - 3.38) | 39 / 8 | 0.63 | (0.20 - 1.97) |
| Ethnicity |  |  |  |  |  |  |
| English or other European | 96 / 35 |  | Ref | 175 / 96 |  | Ref |
| Indigenous | 10 / 2 | 1.06 | (0.10 - 10.92) | 20 / 4 | 0.44 | (0.10 - 1.88) |
| Others or Rather not say | 21 / 4 | 2.30 | (0.38 - 13.87) | 44 / 9 | 0.95 | (0.31 - 2.85) |
| Education |  |  |  |  |  |  |
| Some college or less | 17 / 11 |  | Ref | 37 / 30 |  | Ref |
| College graduate or some university | 41 / 18 | 0.68 | (0.16 - 2.79) | 77 / 33 | 0.54 | (0.22 - 1.32) |
| Bachelor's or higher | 69 / 12 | 0.45 | (0.10 - 2.10) | 125 / 46 | 0.96 | (0.40 - 2.32) |
| Household size |  |  |  |  |  |  |
| Lived alone | 26 / 6 |  | Ref | 42 / 15 |  | Ref |
| Two people | 49 / 20 | 0.22* | (0.04 - 1.30) | 89 / 64 | 0.24*** | (0.09 - 0.65) |
| Three people | 13 / 6 | 1.48 | (0.24 - 9.06) | 34 / 10 | 0.19** | (0.05 - 0.68) |
| Four or more people | 39 / 9 | 0.45 | (0.08 - 2.37) | 74 / 20 | 0.26** | (0.09 - 0.74) |
| **At least 1 household member has any symptom** |  |  |  |  |  |  |
| Yes | 82 / 6 |  | Ref | 149 / 12 |  | Ref |
| No | 19 / 29 | 32.55*** | (8.80 - 120.36) | 48 / 82 | 22.28*** | (10.38 - 47.81) |
| **Smoking** |  |  |  |  |  |  |
| Never smokers or unknown | 79 / 18 |  | Ref | 139 / 51 |  | Ref |
| Current or former smokers | 48 / 23 | 1.89 | (0.60 - 6.02) | 100 / 58 | 1.48 | (0.75 - 2.91) |
| **BMI** |  |  |  |  |  |  |
| Under or normal (< 25 kg/m<sup>2</sup>) | 45 / 11 |  | Ref | 119 / 83 |  | Ref |
| Overweight (25 to 30 kg/m<sup>2</sup>) | 43 / 13 | 0.70 | (0.18 - 2.74) | 111 / 80 | 0.58 | (0.26 - 1.28) |
| Obese (≥ 30 kg/m<sup>2</sup>) | 29 / 13 | 1.09 | (0.28 - 4.22) | 94 / 61 | 0.94 | (0.42 - 2.08) |
| Unknown | 10 / 4 | 1.30 | (0.18 - 9.19) | 24 / 15 | 1.36 | (0.40 - 4.68) |
| **Diabetes** |  |  |  |  |  |  |
| No or unknown | 121 / 38 |  | Ref | 322 / 222 |  | Ref |
| Yes | 6 / 3 | 0.27 | (0.03 - 2.66) | 26 / 17 | 0.51 | (0.16 - 1.67) |
| **Hypertension** |  |  |  |  |  |  |
| No or unknown | 97 / 28 |  | Ref | 262 / 185 |  | Ref |
| Yes | 30 / 13 | 0.61 | (0.17 - 2.14) | 86 / 54 | 0.80 | (0.37 - 1.73) |
| **Constant** |  | 0.11* | (0.01 - 1.14) |  | 0.27* | (0.07 - 1.14) |
| **Observations** |  | 167 |  |  | 347 |  |

Notes:
- a. \*\*\* p<0.01, \*\* p<0.05, \* p<0.1 - b. Analyses exclude 1 person who prefers to self-describe sex

The odds of having no symptoms were lower in larger households (OR = 0.22, 0.04-1.30 among definite seropositives and OR = 0.24, 0.09-0.65 among definite and possible seropositives in households of 2, OR = 0.19, 0.05-0.68 among definite and possible seropositives in households of 3, OR = 0.26, 0.09-0.74 among definite and possible seropositives in households with at least 4 people) compared to those that live alone. However, the odds of having no symptoms were much higher if at least 1 household member also had no symptoms (OR = 32.55, 8.80-120.36 among definite seropositives and OR = 22.28, 10.38-47.81 among definite and possible seropositives).

The odds of having no symptoms among definite seropositives or among definite and possible seropositives did not significantly differ by sex, ethnicity, education, being visible minority, smoking history, BMI, diabetes status, or hypertension status.

##### COVID Symptom Positivity Time Trends

In the sample, 10,575 respondents experienced at least one of the survey symptoms, and 1,191 (6% of the entire cohort) met the study definition of COVID symptom positivity. Appendix Figure 7 shows the proportion of respondents who were COVID symptom-positive in each month among those reporting any symptom, in all ages and by age group. March had the highest proportion of COVID symptom-positive respondents (24.8%) of those reporting any symptom in respondents age 18-39. After March, a slightly higher proportion of respondents age 40-59 were COVID symptom-positive (9.1%) than respondents in the younger age group (7.3% in age 18-39) and in older age groups (4.6% in age 60-69 and 4.4% in age 70 years and older). The counts of COVID symptom positivity among those who experienced any symptom by month and age group are presented in Appendix Table 3.

**Appendix Table 3.** Counts of COVID symptom-positive respondents and household members by age groups and month of symptom onset.

| Age group | COVID symptom-positive | Before March | March | After March | Cannot Recall | Total |
| --- | --- | --- | --- | --- | --- | --- |
| **All ages** | No | 2,604 | 1,840 | 3,309 | 1,631 | 9,384 |
|  | Yes | 351 | 539 | 261 | 40 | 1,191 |
| **18-39** | No | 856 | 736 | 1,420 | 602 | 3,614 |
|  | Yes | 113 | 243 | 112 | 23 | 491 |
| **40-49** | No | 841 | 685 | 1,125 | 531 | 3,182 |
|  | Yes | 140 | 212 | 113 | 11 | 476 |
| **60-69** | No | 582 | 318 | 545 | 324 | 1,769 |
|  | Yes | 77 | 53 | 26 | 2 | 158 |
| **70+** | No | 325 | 101 | 219 | 174 | 819 |
|  | Yes | 21 | 31 | 10 | 4 | 66 |

| Age group | COVID symptom-positive | Before March | March | After March | Cannot Recall | Total |
| --- | --- | --- | --- | --- | --- | --- |
| **All ages** | No | 1,142 | 1,263 | 2,174 | 911 | 5,490 |
|  | Yes | 176 | 355 | 215 | 47 | 793 |
| **18-39** | No | 392 | 552 | 1,022 | 361 | 2,327 |
|  | Yes | 65 | 168 | 95 | 22 | 350 |
| **40-49** | No | 401 | 487 | 777 | 310 | 1,975 |
|  | Yes | 74 | 142 | 89 | 19 | 324 |
| **60-69** | No | 232 | 174 | 287 | 164 | 857 |
|  | Yes | 26 | 34 | 21 | 4 | 85 |
| **70+** | No | 117 | 50 | 88 | 76 | 331 |
|  | Yes | 11 | 11 | 10 | 2 | 34 |

Notes:

In households with at least two household members, the proportion of COVID symptom-positive members (i.e. anyone other than the respondent in the household) was the same as the overall results (6.2%). In these households, 6,283 reported any symptom in someone besides the respondent, of which 1,031 (16%) were COVID symptom-positive. Appendix Figure 8 presents the percent of COVID symptom-positive respondents and/or household members in households with at least two people, by month and respondent’s age group. Of the 1,031 households, 559 reported COVID symptom positivity only in household members and not the respondent, while 472 reported COVID symptom positivity in both household members and the respondent. March was also the peak of COVID symptom positivity for respondents and/or their household members for all age groups, followed by declines of COVID symptom positivity in later months. In March, respondents aged 70 and above were more likely than younger respondents to be the only one in a multi-person household with COVID symptom positivity (17% in age 70+ vs. 12% in age 18-39, 10% in age 40-59, 9% in age 60-69).

**Appendix Figure 8.**
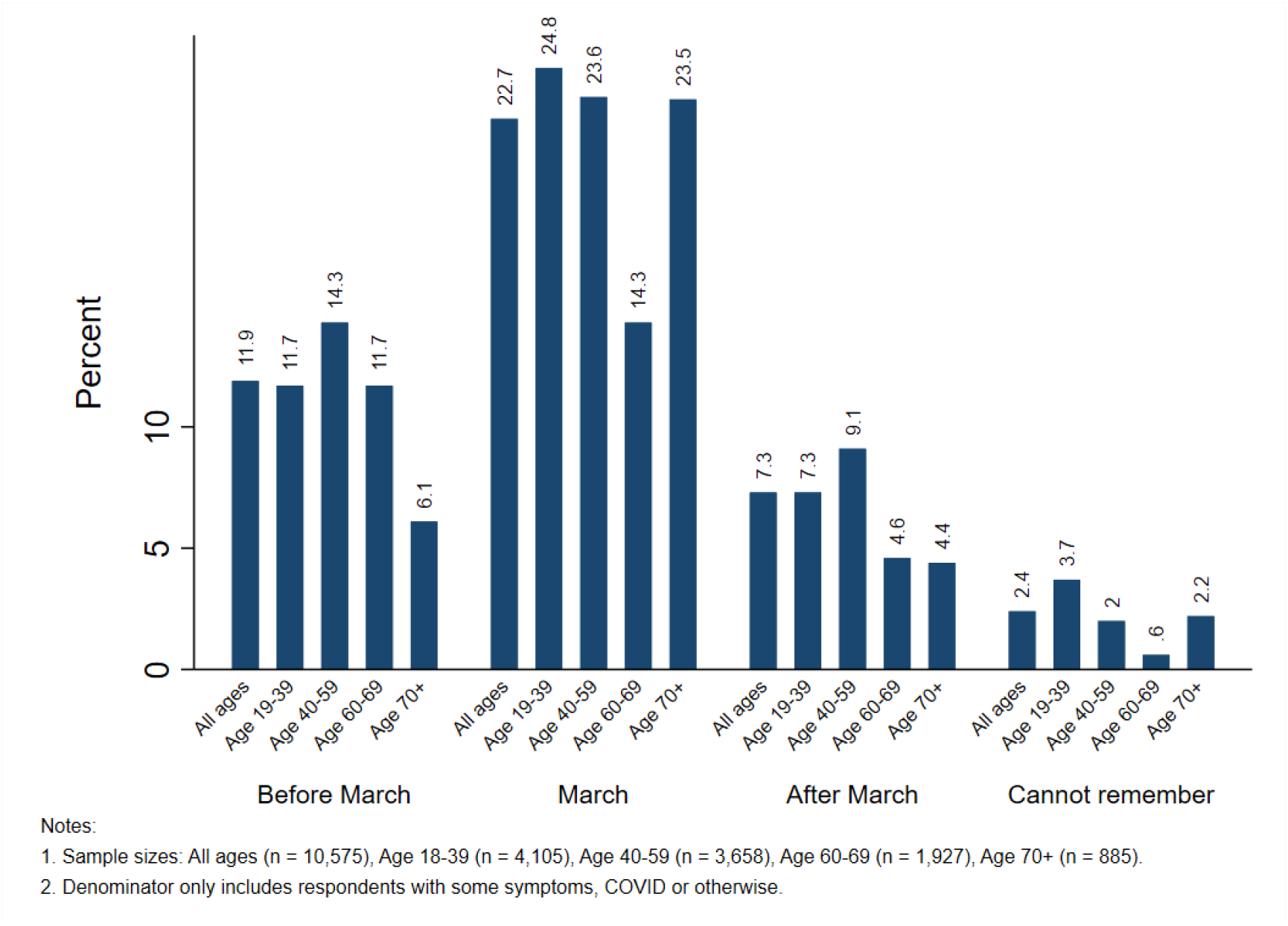
COVID symptom positivity by month of onset and age group.

### Section 4: Mortality Data Sources and Calculations

We tracked COVID mortality reporting from the Public Health Agency of Canada,^15^ Statistics Canada,^16^ and other sources including cross-checks against media and daily public health reporting of COVID deaths, which have continued on an emergency basis. This showed that the peak of mortality occurred in May and June, following the peaks of when PCR-based SARS-CoV-2 infection was diagnosed (April) and the peak of symptoms (March; Appendix Figure 9, data shown for March through May).^17^

**Appendix Figure 9.**
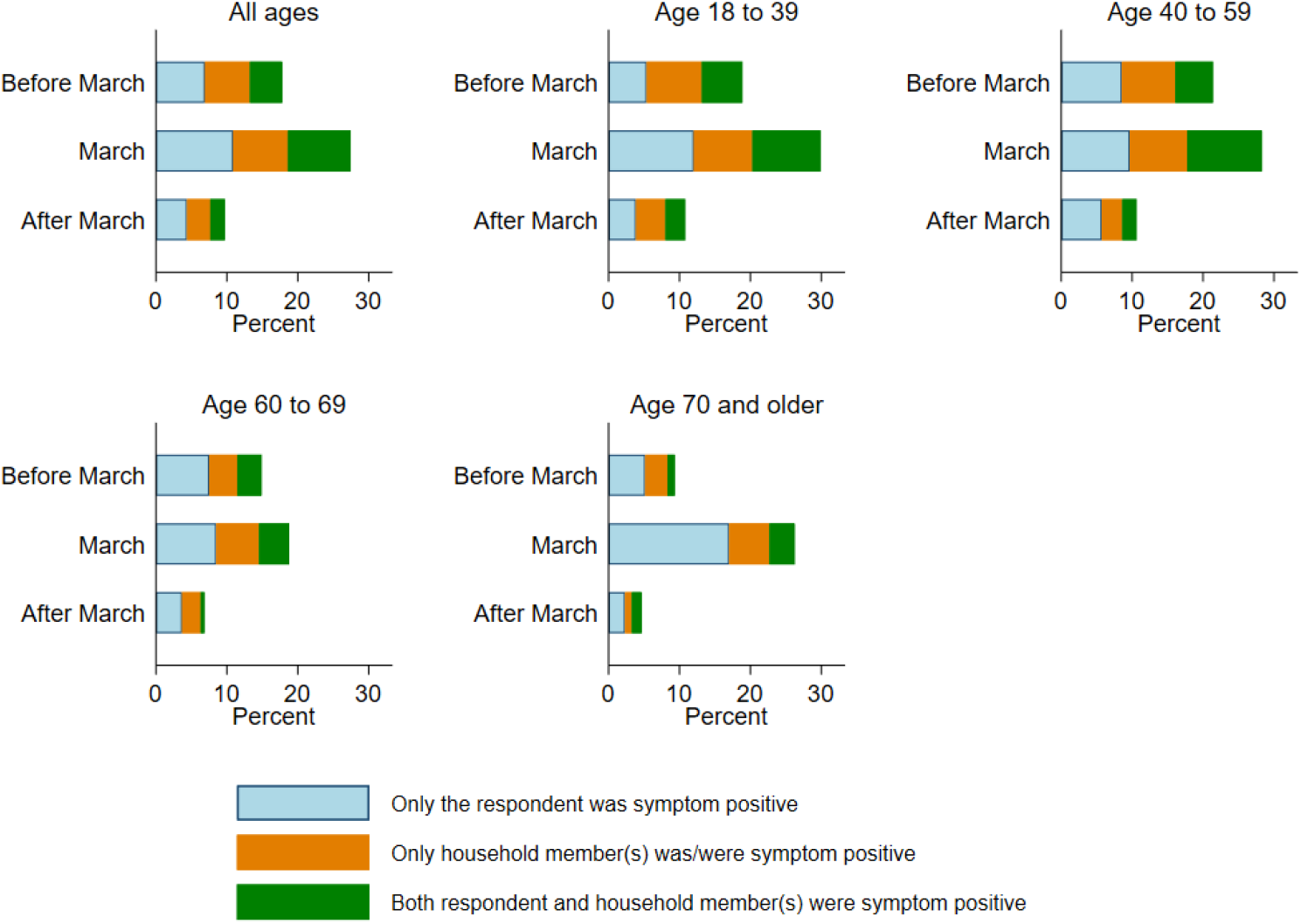
Percent of COVID symptom-positive respondents and/or household member(s) by time and respondent age group.

**Appendix Figure 10.**
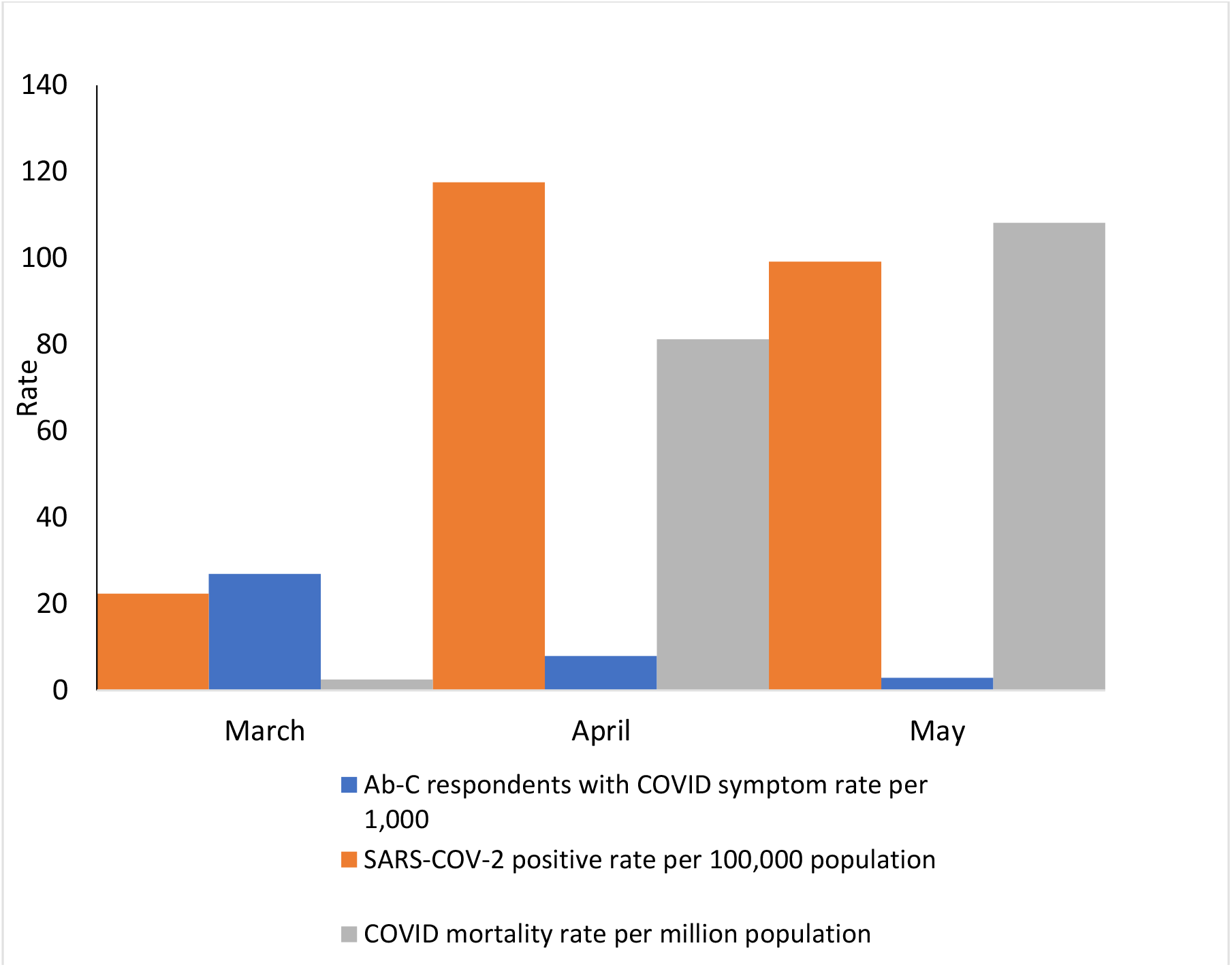
COVID symptom positivity among Ab-C respondents compared to COVID cases and deaths in Canada.

We documented 9,045 cumulative COVID deaths in Canada as of Sept 1, 2020. We allocated these to age groups, keeping nursing home deaths as their own category, regardless of the age and sex of decedents. Deaths before age 50 rarely occur in nursing homes, so we directly recorded the few deaths before age 50 into the analyses, as no parsing by nursing home deaths was needed. Next, we allocated deaths above age 50 to nursing home or non-nursing home deaths using a Bayesian model. The proportion of COVID-19 deaths in long-term care homes was computed using the model below implemented in the Stan software.^18^

Write *Y*_1*ij*_ and *Y*_2*ij*_ as the (unknown) number of deaths in nursing homes and otherwise, respectively, for age/sex group *i* in region *j*. Observed are the number of deaths in nursing homes *Y*_1·*j*_ summed by age/sex groups and age-sex specific deaths *Y*_*·ij*_ including both deaths in nursing homes and not in nursing homes by group and region. Using *P*_*kij*_ and *λ* _*kij*_ to refer to the census-obtained populations and expected counts, *θ*_*i*_ to refer to age-sex specific log relative risks, and *γ*_1*j*_ and *γ*_2*j*_ to indicate region-specific log relative risks for residents in nursing homes and non-nursing home residents, the model used is as follows:

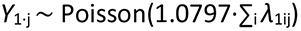

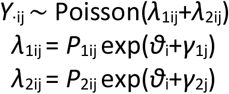

The value 1.0797 is a correction applied because the total number of COVID deaths reported in the dataset on deaths in nursing homes exceeded the total in the age-sex specific dataset by roughly 8%. The key feature of this model is it does not allow for an interaction between age and whether individuals live in nursing homes. The increase in risk resulting from living in nursing homes is the same, proportionately, for individuals in every age-sex group. This assumption permits inference to be made on the age-sex distribution of deaths in nursing homes on the basis of the aggregated data made public. Writing *ρ*_*ij*_ as the proportion of deaths in nursing homes for group *i* of region *j*, the number of deaths in nursing homes can be sampled as

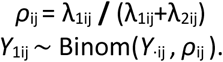

Uninformative prior distributions were used for all model parameters.

This procedure allowed us to allocate 7,009 COVID deaths to nursing homes (and this proportion of 77% of total COVID deaths was consistent with various other analyses and media reports^19^). The remaining deaths at older ages (50 or higher) occurred outside of nursing homes and were allocated to their respective age groups as in Table 2.

Next, we calculated the excess deaths in nursing homes, based on documented deaths in prior, non-COVID years. The Statistics Canada 2019 mortality data provide location of death for nearly all provinces (Appendix Table 4). Nursing home deaths are typically recorded as in “Other health care facility,” although there are some variations (such as some nursing home deaths in Manitoba appearing as hospital deaths). Direct data for Quebec were not available, so as an approximation, Statistics Canada ran a logistic regression model of hospital vs. other health care facility fit to the 2017 to 2019 national data (less Quebec and those deaths in Manitoba that occurred prior to August 2018).^20^ Covariates were sex, age, and underlying cause of death, using the broad age and cause categories used in monthly releases on excess mortality. The Quebec data were scored using the model parameters, and the predicted probabilities were used to impute location of death where the distinction between hospital and other health care facility was not made.

**Appendix Table 4.** Location of deaths in 2019, in Canada

| Province of death occurrence | Hospital | Private dwelling | Other health care facility | Other or unknown locality | Total | % of deaths in other health care facility |
| --- | --- | --- | --- | --- | --- | --- |
| Ontario | 58,185 | 20,965 | 20,265 | 7,895 | 107,310 | 19% |
| Quebec | 37,085 | 8,165 | 15,955 | 5,475 | 66,680 | 24% |
| **British Colombia** | 16,415 | 7,365 | 13,530 | 1,250 | 38,560 | 35% |
| **Alberta** | 13,880 | 4,755 | 4,660 | 2,900 | 26,195 | 18% |
| **Manitoba** | 6,830 | 1,030 | 2,450 | 885 | 11,195 | 22% |
| **Nova Scotia** | 5,680 | 1,875 | 1,980 | 315 | 9,850 | 20% |
| **Saskatchewan** | 5,070 | - | 60 | 4,420 | 9,550 | 1% |
| **New Brunswick** | 4,505 | 1,100 | 1,800 | 385 | 7,790 | 23% |
| **Newfoundland** | 2,980 | 885 | 1,260 | 105 | 5,230 | 24% |
| **Prince Edward Island** | 735 | 170 | 420 | 25 | 1,350 | 31% |
| **Northwest Territories** | 95 | 55 | 40 | 15 | 205 | 20% |
| **Nunavut** | 45 | 60 | 25 | 35 | 165 | 15% |
| **Total** | 151,505 | 46,425 | 62,445 | 23,705 | 284,080 | 22% |

This analysis yielded a total of 62,445 nursing home deaths annually in the pre-COVID era. This is from a total denominator, measured in 2016, suggesting a mid-year nursing home population of 168,205 with another 86,145 living in facilities that are a mix of both a nursing home and a residence for senior citizens.^21^ To specifically estimate the mid-year Canadian nursing home population in 2016, we noted using known populations in Ontario for those living in a nursing home vs. a residence for senior citizens that approximately 50% or 43,073 of the 86,145 persons across Canada living in facilities that are a mix of both a nursing home and a residence for senior citizens were likely living specifically in nursing homes. We further understand that the nursing home population across Canada has not significantly changed in the last 5 years. Therefore, we estimate that in 2020, the mid-year nursing home resident population in Canada was 211,277 persons. We note that this number is likely a more conservative value, as Canadian nursing homes tend to run at near 100% capacity, so that the number of actual nursing home residents being supported in the approximate 211,277 Canadian nursing home beds in a given year could be as high as 253,532 Canadians.

Drawing upon methods published earlier for HIV,^22^ we estimated the relation of the SARS-CoV-2 prevalence (*P*) to COVID-attributable mortality using a simplified approach. First, we used the formula *P* = ((M / IFR)/N), where IFR is infection fatality rate). M in this case is the 7,009 COVID deaths in nursing homes, and N is the total nursing home population. There is no reliable measure of SARS-CoV-2 infection in nursing homes in Canada, so we relied on the upper and lower confidence intervals from a recent empiric analysis of IFRs^22^ for nursing homes in France, where the IFR was 22.5% (95% CI 19.1-25.8%). Applying these two IFRs to the Canadian nursing home deaths and population yields a prevalence of infection in nursing homes of about 13-17%, which is consistent with reviews of excess mortality (annualized for the short duration of COVID in the first viral wave^23^ and reviews of nursing home deaths).^25^ The estimated IFR in France has a lower and higher value (11.71% [95% CI 10.03-13.55] and 35.13% [95% CI 30.10-40.64], respectively). Applying the most extreme 95% CIs yields 9-32% prevalence (data not shown) as the proportion of COVID deaths in Canadian nursing homes.

